# Cell-type specific and multiscale dynamics of human focal seizures in limbic structures

**DOI:** 10.1101/2023.03.06.23286778

**Authors:** Alexander H. Agopyan-Miu, Edward M. Merricks, Elliot H. Smith, Guy M. McKhann, Sameer A. Sheth, Neil A. Feldstein, Andrew J. Trevelyan, Catherine A. Schevon

## Abstract

The relationship between clinically accessible epileptic biomarkers and neuronal activity underlying the seizure transition is complex, potentially leading to imprecise delineation of epileptogenic brain areas. In particular, the pattern of interneuronal firing at seizure onset remains under debate, with some studies demonstrating increased firing while others suggest reductions. Previous study of neocortical sites suggests that seizure recruitment occurs upon failure of inhibition, with intact feedforward inhibition in non-recruited territories. We investigated whether the same principles applied also in limbic structures.

We analyzed simultaneous ECoG and neuronal recordings during 34 seizures in a cohort of 19 patients (10 male, 9 female) undergoing surgical evaluation for pharmacoresistant focal epilepsy. A clustering approach with five quantitative metrics computed from ECoG and multiunit data was used to distinguish three types of site-specific activity patterns during seizures, at times co-existing within seizures. 156 single-units were isolated, subclassified by cell-type, and tracked through the seizure using our previously published methods to account for impacts of increased noise and single-unit waveshape changes caused by seizures.

One cluster was closely associated with clinically defined seizure onset or spread. Entrainment of high-gamma activity to low-frequency ictal rhythms was the only metric that reliably identified this cluster at the level of individual seizures (*p* < 0.001). A second cluster demonstrated multi-unit characteristics resembling those in the first cluster, without concomitant high-gamma entrainment, suggesting feedforward effects from the seizure. The last cluster captured regions apparently unaffected by the ongoing seizure. Across all territories, the majority of both excitatory and inhibitory neurons reduced (69.2%) or ceased firing (21.8%). Transient increases in interneuronal firing rates were rare (13.5%) but showed evidence of intact feedforward inhibition with maximal firing rate increases and waveshape deformations in territories not fully recruited but showing feedforward activity from the seizure, and a shift to burst-firing in seizure-recruited territories (*p* = 0.014).

This study provides evidence for entrained high gamma activity as an accurate biomarker of ictal recruitment in limbic structures. However, our results of reduced neuronal firing suggest preserved inhibition in mesial temporal structures despite simultaneous indicators of seizure recruitment, in contrast to the inhibitory collapse scenario documented in neocortex. Further study is needed to determine if this activity is ubiquitous to hippocampal seizures or if it indicates a “seizure-responsive” state in which the hippocampus is not the primary driver. If the latter, distinguishing such cases may help refine surgical treatment of mesial temporal lobe epilepsy.

## Introduction

Understanding the neuronal dynamics of seizure genesis and spread are important for efforts to improve medical and surgical management of epilepsy. While surface and intracranial EEG can provide valuable localizing information, they are relatively insensitive to neuronal firing activity; indeed, similar appearing EEG features may be associated with very different patterns of unit activity. Previously, we described a dual-territory seizure structure, in which a core of intense neural firing that drives the seizure can be distinguished from the relatively large “penumbra” in which neural firing is inhibited^1–8^. This is consistent with observations of intense, synchronized neuronal firing following the ictal transition from *in vitro* studies^6,9–11^. Conversely, others have proposed that seizures arise from the aggregate activity of pathological diffuse networks^12–14^, based on observations of modest, heterogenous firing rate changes^15–22^. However, our prior work was limited due to relatively small study populations with a variety of epilepsy syndromes, and being confined to neocortical onsets, which may show different cellular activity to the hippocampal seizures that have been the focus of much prior animal and human work^20,22–27^.

A significant issue affecting all such studies is the lack of a gold standard for identifying the cellular activity that defines the ictal transition. Attention has focused on the role of fast-spiking interneurons, with animal studies and a recent study in human mesial temporal structures demonstrating increased fast-spiking interneuron firing rates prior to the ictal transition^28–30^, and activating these cells optogenetically can induce seizure-like events *in vitro*^31,32^. An independent human study, however, demonstrated decreased interneuronal firing rates just prior to hippocampal ictal invasion from a neocortical source^33^. We hypothesized that the observed early ictal interneuronal firing patterns depend on the nature of local seizure involvement, which can be characterized by aggregate EEG and neuronal firing patterns. Here, we utilized multi-scale neuronal activity patterns, recorded using stereo-electroencephalography (sEEG) incorporating microwires, in patients undergoing surgical evaluation for pharmacoresistant epilepsy, to distinguish different ictal territories. In this way, we can identify cell-type specific neuronal firing patterns within a well-defined clinical and spatial context.

## Materials and methods

### Human recordings

Patients undergoing stereotactic depth electrode placement for presurgical evaluation of intractable focal epilepsy were simultaneously implanted with “Behnke-Fried” type micro-macro arrays (Ad-Tech Medical, Racine WI). Research was conducted under the oversight of Columbia University’s Institutional Review Board; all patients provided informed consent prior to implant. Recording details are as described in our prior publication^3^. Electrodes were localized using iELVis with co-registered postimplant CT and preimplant T1 volumetric MRI scans^34^. Clinical determinations of the epileptogenic zone (EZ), consisting of the seizure onset zone (SOZ) and path of spread, as well as seizure semiology classification, were made by treating physicians and confirmed by a qualified epileptologist (C.A.S.) prior to analysis.

### Data processing

Processing and analyses were performed using custom code written in MATLAB (MathWorks, Natick MA). Analysis focused on microwires and their nearest macroelectrode (“macro-micro pair”). Microwire and macroelectrode signals were de-meaned, then common average re-referenced. The macroelectrode signal was processed into two data streams using symmetric 512^th^- order FIR bandpass filters: a low frequency (2–20 Hz) band capturing the dominant ictal rhythm and a high-gamma band (80–150 Hz) as a proxy for population firing^35–37^. To quantify interictal high frequency oscillation (HFO) rates for each macro-micro pair, HFOs were detected from 10-minute sleep epochs using RIPPLELAB^38^ with visual review to exclude false positives, following published guidelines^39^. Multi- and single-unit activity were isolated from the 300Hz–5kHz band from each microwire recording as previously described^3^. Briefly, after artifact removal, extracellular action potentials were detected and clustered into single-units in the 5-minute pre-ictal period. Units were then tracked through the seizure using a probabilistic template-matching algorithm^3^, to minimize loss of units due to obscured clustering or waveshape deformation that could erroneously give the impression of reductions in firing rates. Only macro-micro pairs with isolatable single-unit activity were included.

Analyses focused on two epochs: a 5-minute period prior to seizure onset (“pre-ictal”) and an “ictal” period from seizure onset until the start of either a pre-termination pattern as described previously^35^ or, in the case of focal to bilateral tonic-clonic seizures, the time of transition to tonic-clonic semiology (identified by expert review of video EEG [C.A.S.]). Both the tonic-clonic phase and pre-termination stage—characterized by irregular inter-discharge intervals and reduced per-discharge high-gamma activity—feature unique spatiotemporal dynamics^35,40–42^ and were excluded from analysis.

### Neuronal activity metrics

In order to assess seizure-related neural patterns, we used metrics designed to capture relevant multiscale features from each macro-micro pair and seizure. Up to three electrographic seizures with units trackable across the ictal transition were studied (**Supplementary Table 1**). The following metrics were utilized: multi-unit firing rate (**FR**); multi-unit firing entrainment to the low frequency rhythm (**MU-E**); unit waveform width (“full width at half maximum”; **FWHM**); instantaneous macroelectrode high-gamma (80–150 Hz) amplitude (**HGA**); and high-gamma phase-amplitude coupling to the low frequency rhythm (**PLV**_**HG**_; **Fig. 1**). FR was calculated as the ratio of the ictal and pre-ictal firing rates. MU-E was calculated by extracting the instantaneous phase of the low-frequency signal at each multi-unit spike time, and calculating the Rayleigh Z-statistic. FWHM was calculated as the ratio of the average ictal multi-unit FWHM to the average pre-ictal multi-unit FWHM for each microwire^3^. HGA was determined using the envelope of the Hilbert transform on the high-gamma filtered signal from the microcontact and normalized to the mean pre-ictal value. Phase-locking value (PLV_HG_) was then computed from the Hilbert-transformed high-gamma signal to the phase of the low-frequency rhythm using 3-second sliding windows advancing in 333 millisecond increments^43–45^, then averaged for the ictal epoch.

**Fig. 1.**
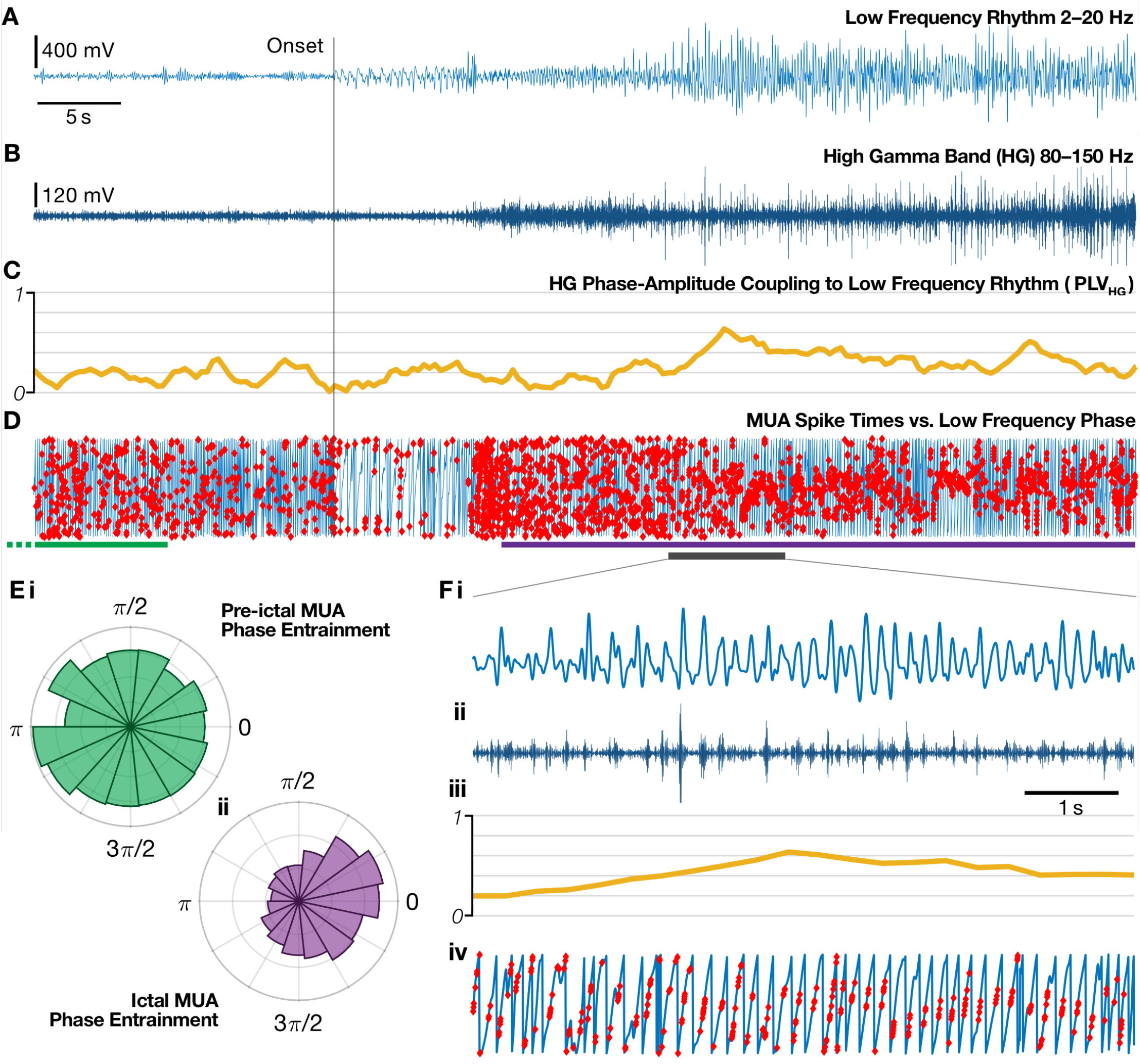
Illustration of neural activity metrics. The raw signal obtained from the clinical macroelectrode is filtered into two bands: the 2–20 Hz low frequency rhythm (**A**) and the 80–150 Hz high-gamma band (**B**). **C**. The coupling between the high-gamma amplitude and the low frequency phase (PLVHG) can then be calculated across these signals. **D**. Detected multi-unit spikes from the microwires (red dots) provide firing rate and multi-unit entrainment (MU-E) to the phase of the low frequency rhythm (blue line). **E**. Polar histograms for the phases of all multi-unit spikes from the regions marked in D, comprising the same number of spikes in each of the pre-ictal epoch (**i**, green, starting prior to region plotted) and the ictal epoch (**ii**, purple), with corresponding Rayleigh z-statistic values of 2.90 and 107.3 respectively. **F**. Expanded section from panels A–D as marked by the grey bar.

### Dimensionality reduction and electrode cluster analysis

To ensure metrics across all macro-micro pairs approximated a normal distribution for comparative purposes, each was Box-Cox transformed^46^ prior to subsequent analysis in feature space. t-distributed stochastic neighbor embedding (**t-SNE**) was used to visually assess groupings among macro-micro pairs. Principal component (**PC**) analysis was then used to validate the presence of clusters in the dataset. Components that explained >99% of the data were retained for cluster analysis.

To determine the optimal number of clusters in PC space, successive *k*-means solutions were calculated and evaluated using the Calinski-Harabasz index. Solutions with Calinski-Harabasz values with <1% probability of being observed in a null dataset, created by randomly shuffling the distances between observations in PC space and recalculating the Calinski-Harabasz index over 1000 runs, were also evaluated for cluster stability (Jaccard coefficient > 0.6)^47^ using bootstrap resampling (R “fpc” package clusterboot function)^48,49^.

The relative contribution of each metric to PC clustering was calculated by summing the product of the variance explained for a given PC by the metric coefficients for that PC, then dividing the result by the sum of coefficients for a given metric across all PCs. To visualize each metric’s contribution to the clustering, a given metric was removed from the dataset and PCA was re-run on the remaining data but with each macro-micro pair retaining its original cluster ID. Cluster center separation was calculated via Hotelling’s two-sample T^2^ test on Mahalanobis distances, with a Bonferroni-Holm-corrected F-test to determine statistical significance^50^.

### Single metric comparisons

The original, non-Box-Cox transformed data for each metric was compared between clusters using a two-tailed Wilcoxon rank-sum test with Bonferroni-Holm correction for multiple comparisons. Line-length served as a quantitative proxy for the clinically determined EZ^51,52^. The interictal HFO rates between clusters was compared using a two-sample t-test.

### Single-unit putative cell-type classification

Single-units were sub-classified into putative fast-spiking (FS) and regular-spiking cell-types based on waveform shapes and cell-intrinsic metrics. For each unit, the mean waveform was calculated as the spike-triggered average from the raw, unfiltered original signal, z-scored and a 2-component Gaussian mixture model was fitted to the z-scored waveform’s scores in PC space, which has been shown to separate FS interneurons from the regular spiking (RS) population^53^. FS cells primarily represent PV^+^ interneurons due to their fast hyperpolarization via Kv3.1/Kv3.2 channels^54,55^, i.e. roughly 40% of interneurons^56^, resulting in the remaining RS cells comprising a mixture of predominantly excitatory and a minority of inhibitory cells. To extract putative inhibitory cells from the RS population, the autocorrelation for each unit that was not already classified as a FS interneuron was fitted with a set of exponentials using the “fit_ACG” function from “CellExplorer”^57^. The τrise exponential fit captures the timing of the rise in probability of following action potentials occurring after an initial spike from that neuron, a feature which can separate pyramidal cells from non-FS interneurons, due to their cell-intrinsic firing dynamics^57,58^. We fitted a 2-component Gaussian mixture model to the τrise values for each RS unit, providing probabilities for the subclassification of each RS cell as inhibitory (“RS interneuron”) or excitatory.

To assess the expected proportion of action potentials that may have been subthreshold at any given moment, the distribution of the unit’s voltage at detection was fitted with a Gaussian^59^, and to account for any drift, that Gaussian was shifted such that its mean was equal to the smoothed spline at any given time point. The cumulative probability of this shifted Gaussian that was below the original threshold used for spike detection provides an estimate of the rate of missing spikes for each unit. Cessations of firing were calculated as the moment the ictal firing rate dropped to zero or < -3 SD of the pre-ictal rate, whichever was lower (in order to prevent low-firing-rate units from the pre-ictal epoch that reduced to zero by chance being deemed cessations), and their probabilistic firing rate had to remain confident that no action potentials occurred for the rest of the seizure (i.e. < 50% chance of a single spike continuously).

For statistical analyses of single-unit firing patterns, a KS-test for normality was calculated for the comparison groups, and if all were normally-distributed an unpaired *t*-test was used, otherwise the non-parametric Mann-Whitney U test was used.

### Data availability

The data that support the findings of this study are available upon reasonable request from the corresponding author. The data are not publicly available to protect the privacy of research participants.

## Results

Recordings from 34 seizures across 19 patients had isolatable single-units in the peri-ictal period (**Supplementary Tables 1&2**; 10 female; 19–55 years old), with 156 single-units across 52 microwire arrays total, averaging 2.7 arrays per patient (range 1–4).

### Clustering of ictal neuronal activity metrics demonstrates up to three distinct patterns

Neural activity metrics during seizures were computed for macro-micro pairs, then Box-Cox transformed for visualization (see Methods). Each seizure may be represented by multiple macro-micro pairs, i.e. multiple recording locations (**Supplementary Table 2**), and each macro-micro pair was treated as its own entity for analysis of activity patterns. Dimensionality reduction with t-distributed stochastic neighbor embedding (t-SNE) revealed the visual presence of two to three robust groups (**Fig. 2A**). However, since t-SNE is not in itself a reliable clustering method^60^, principal component analysis (PCA) with *k*-means clustering was used on the Box-Cox transformed data to analyze the observed groupings. The first four principal components (PCs) explained >99% of the variance within the data and all subsequent analyses were conducted across these components.

**Fig. 2.**
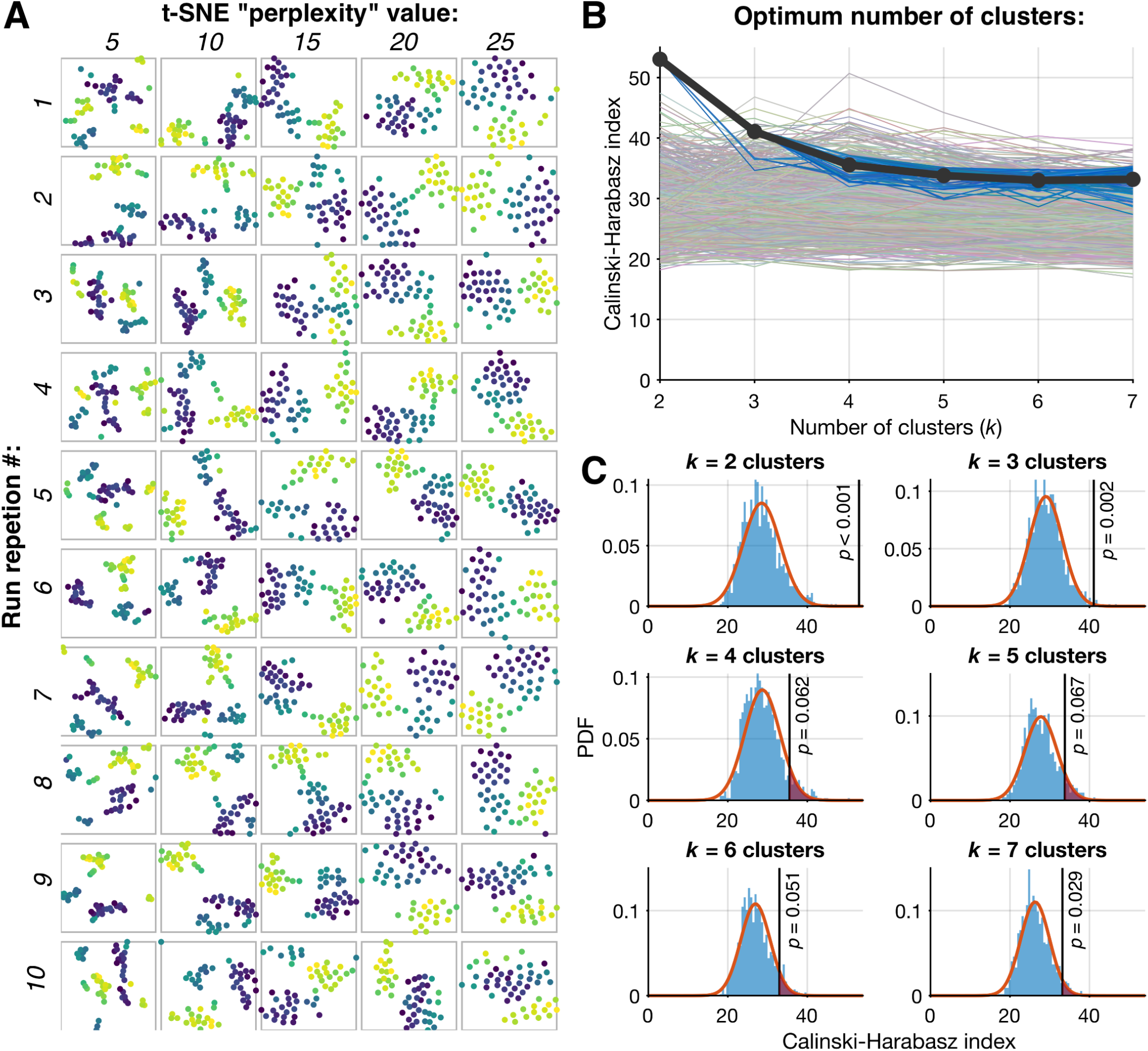
Distinct neural activity patterns identified with clustering techniques. **A**.Each macro-micro pair’s metrics plotted in feature space after dimensionality reduction (t-SNE), revealing 2 to 3 visually distinguishable clusters across multiple runs and different “perplexity” values. For ease of interpretation, each datapoint is color-coded based on the results of an average run (run 6; perplexity = 15). **B**. To derive null distributions for Calinski-Harabasz (CH) indices that would arise by chance, principal component scores between data points were randomly shuffled and the CH index recalculated 1000 times (faded multi-colored lines in background). The CH index is then calculated on the original data 100 independent times (blue lines) and the final CH index is calculated as the mean of those runs (thick black line). **C**. Gaussian curves are fitted to the null distributions found in B, (fit in orange, histogram of shuffled data in blue), from which the probability of finding a value as extreme as the observed CH index can be calculated (black line). Solutions with k > 7 produced clusters comprising single members. While k = 7 was unlikely to arise by chance, the solution was unstable (see Methods; 7-cluster mean Jaccard values range = 0.561–0.836).

The PCA results were found quantitatively to contain more than 1 cluster (p < 0.001; Duda-Hart^61^ test). The optimal number of clusters was calculated by running successive *k*-means solutions and comparing their Calinski-Harabasz (CH) index to null distributions (see Methods). Solutions that were likely to have arisen by chance (*k*=4– 6), were unstable as assessed by mean Jaccard coefficients^47^ (*k*=7), or produced clusters with only single members (*k*>7), were excluded from further analysis. The remaining 2- and 3-cluster solutions were both stable and unlikely to have arisen by chance (**Fig. 2B&C**). The 2-cluster solution was considered the optimal solution as it maximized the CH index^62^ (**Fig. 2B**), while the 3-cluster solution identified a subcluster of macro-micro pairs within one cluster of the 2-cluster solution (explored in detail later).

We sought to characterize each metric’s contribution to the optimal clustering seen in PC space, in order to quantify its potential for localization. PLV_HG_ explained 45.66% of the variance across the first four PCs, followed by MU-E, FR, HGA, and FWHM (21.40%, 19.55%, 10.01%, and 3.11% respectively). To assess for the robustness of the findings and ensure that the 2-cluster separation was not due to any one feature, we re-ran PCA using a leave-one-out analysis, while maintaining cluster IDs from the original 2-cluster solution: cluster center separation remained significant in each metric’s absence (p < 0.05 in each, Bonferroni-Holm-corrected F-test; F-value for each: PLVHG-excluded = 5.08; HGA-excluded = 105.61; FWHM-excluded = 105.60; FR-excluded = 94.13; MU-E-excluded = 107.92). Accordingly, the strongest metrics for grouping the data were those based on entrainment of higher frequency activity to the ictal rhythm: PLV_HG_ followed by MU-E, more so than their underlying firing rate or signal amplitude without respect to the ictal rhythm.

### Distinguishing activity patterns of recruited tissue

A single cluster, identical across both solutions, closely resembled neuronal firing patterns previously associated with the ictal core^1,3,5,43^ (**Figs. 3A & 5A**), so we hypothesized this cluster represented recruitment, labeling it “**R**”. This cluster was comprised entirely of 95% (19/20) of the contacts in the clinically-determined epileptic zone (EZ; p < 10^−4^, Fisher’s Exact Test; 74% (14/19) in SOZ, 26% (5/19) in the clinically-determined path of spread). The remaining cluster was labeled non-recruited (**NR**; sub-grouped as **NR1 & NR2** in the derivative 3-cluster solution).

**Fig. 3.**
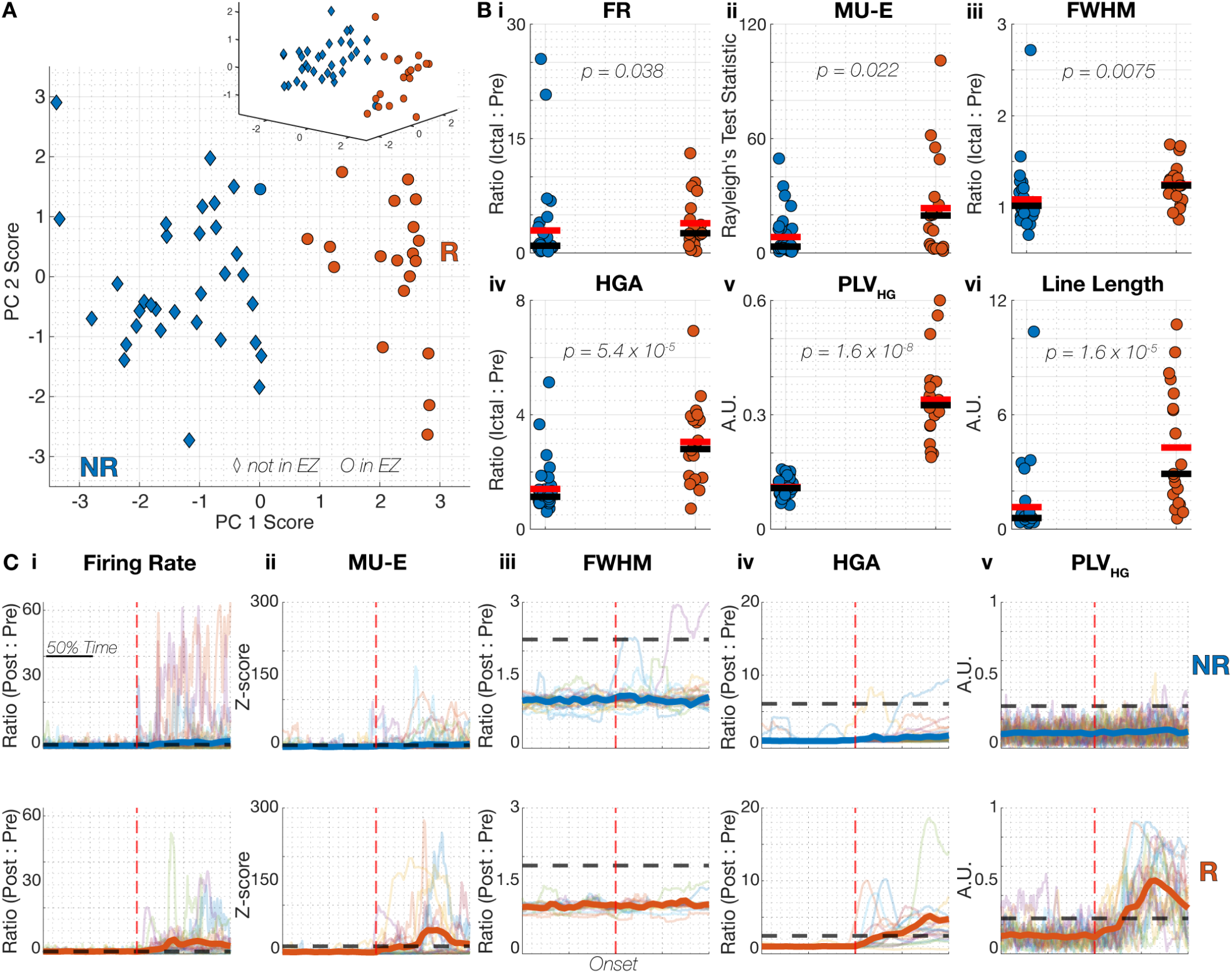
Ictal activity patterns revealed by the 2-cluster solution. **A**.The largest principal components are plotted in 2D (inset in 3D), color coded as the non-recruited group, “NR” (blue), and recruited group, “R” (orange-red); almost all of the clinically determined EZ sites correspond to the recruited group. **B**. Box plots comparing each metric by group. Line-length is shown as a quantitative proxy for the clinically determined EZ and was not included in the dataset used for principal component analysis. Note that while all metrics showed significant differences between recruited and non-recruited groups, complete separation of the groups was apparent only for the PLVHG metric (**Bv**). Group average and median are represented by the red and black bars, respectively **C**. The trajectory for each macro-micro recording in each metric plotted against normalized time (faded lines in background), and aligned to seizure onset (red dashed line). The thick blue and orange-red lines represent the mean value of each metric through time for the non-recruited and recruited group, respectively. The black-dotted line represents +2.5 SDs above the pre-ictal mean for each metric. Note how consistently PLVHG changes across the ictal transition for each member in their respective groups (**Cv**): by about halfway through the ictal period (“50% time”) the average PLVHG across members in the recruited group was more than 2.5 SDs above the preictal average, whereas ictal PLVHG is unchanged from pre-ictal baseline for each member in the non-recruited group.

Macro-micro pairs in R demonstrated larger values compared to pairs in the NR cluster for all 5 metrics used in the cluster analysis (**Fig. 3B**; R>NR, Bonferroni-Holm rank-sum p < 0.05 in each), consistent with predicted characteristics of recruited tissue^1–5^. Line-length, a control metric as a proxy for visual EEG but not used in clustering, was similarly higher in R (**Fig. 3B**; p < 0.05, Bonferroni-Holm rank-sum).

To assess this designation independently from the data used for clustering we contrasted interictal HFO rates between groups using semi-automated HFO detection on 10-minute interictal sleep recordings, given the association between HFOs and seizure source regions^38,63,64^. HFOs (80–600 Hz) were detected in 11% of macroelectrodes (2/18) in NR versus 71% (10/14) in R (*p* = 0.00047, chi-square test), all in hippocampal electrodes. Greater rates were found in contacts from R compared to NR both for HFOs (13.3 ± 16.1 HFOs/10min vs. 0.8 ± 3 HFOs/10min; p = 0.016, unpaired t-test) and fast ripples (5.8 ± 8.0 fast ripples/10min vs. 0.2 ± 0.7 fast ripples/10min; p = 0.027, unpaired t-test). Despite the overall group differences, there was considerable overlap in most metrics—only phase-locking of high-gamma activity to the ictal rhythm (PLVHG) was able to discriminate the two groups without overlap between their distributions (**Fig. 3Bv**). Similarly, PLVHG displayed temporal stereotypy during each seizure (**Fig. 3C**), indicating an evolution from non-recruited to recruited status at these territories. To further explore this, we fitted a Gaussian mixture model to the two metrics accessible to routine clinical recordings (PLVHG and HGA), in order to derive instantaneous probabilities of each location being recruited at any given time (**Fig. 4**). Doing so revealed that recruitment always arose from NR feature-space (**Fig. 4B-F**), and followed stereotyped paths across seizures prior to recruitment (**Fig. 4D**) and once recruited (**Fig. 4E&F**).

**Fig. 4.**
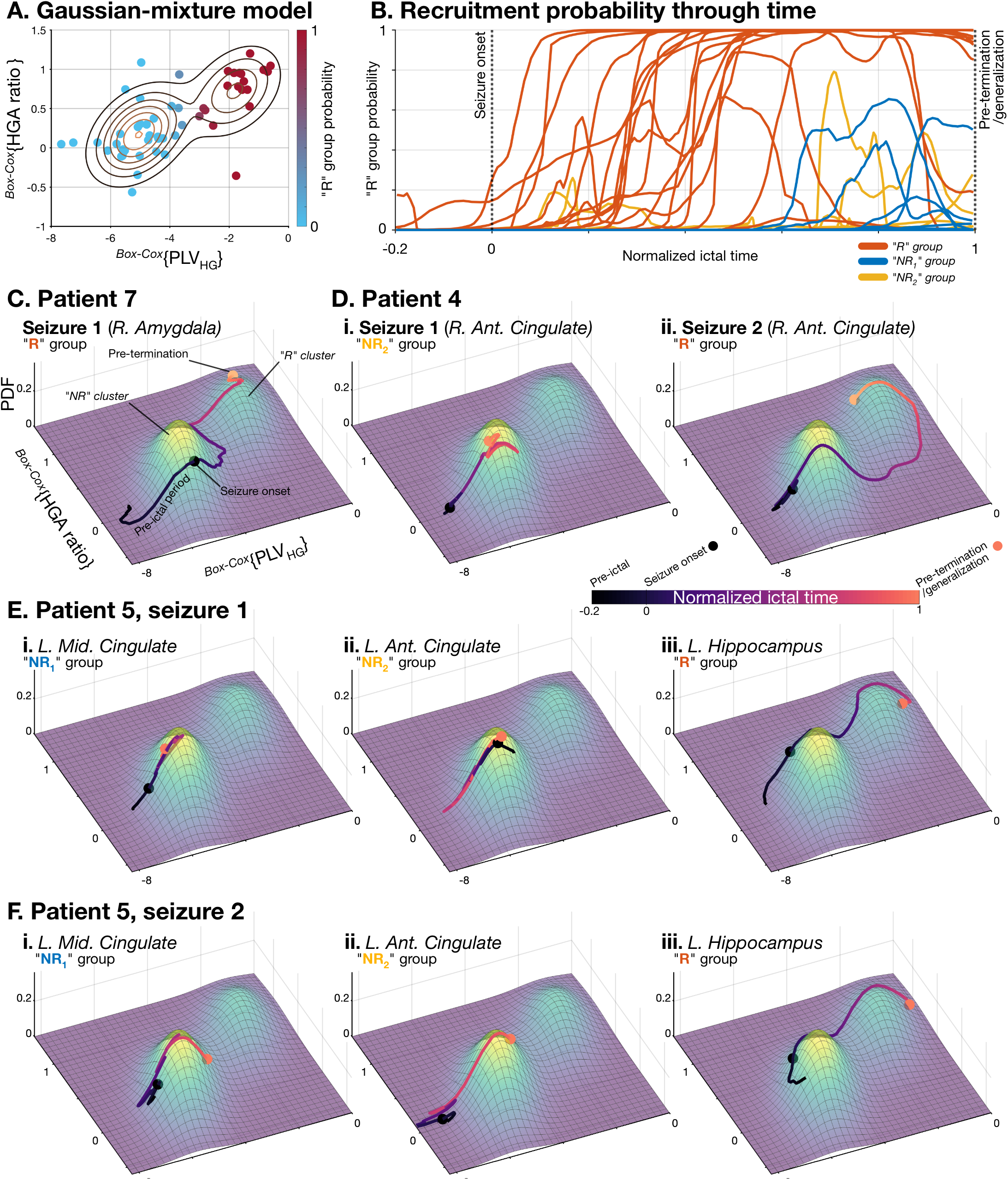
Recruitment follows a trajectory starting in the non-recruited feature space. **A**.Fitting a Gaussian mixture model to the Box-Cox transformed macro-electrode metrics produces a stable feature space that can provide probabilities of belonging to the recruited vs. non-recruited groups across any given value. **B**. The instantaneous probability of being in recruited tissue for each electrode, throughout the pre-ictal and ictal epoch (normalized time), calculated by transforming the continuous PLVHG and HGA values onto the Box-Cox feature space using the lambda values from the original clustering. Traces are color-coded by original clustering group (“R” in orange, “NR1” in blue, “NR2”in yellow), showing the rise in probability that the tissue recorded in the “R” group has transitioned to recruited as the seizure progresses, arising from a non-recruited state. **C**. Example trajectory of a single electrode and seizure in the feature space from A, color coded by normalized ictal time, with seizure onset and pre-termination marked with circles. Heatmap below shows the probability density function, revealing the dense regions for “NR” and “R” values. This electrode was in the “R” group for this seizure, and it can be seen to start in the non-recruited region, and progress to the feature space corresponding to recruitment before pre-termination. **D**. Same layout as C, showing a single electrode that remained unrecruited in seizure 1 (**i**, focal unaware seizure) and then became recruited in seizure 2 (**ii**, FTBTC seizure). Note the nearly identical trajectory in seizure 2 prior to transitioning to recruitment. **E & F**. Three paired electrodes from two seizures in a single patient, showing stereotypy of trajectory across seizures within each cluster: (**i**) a mid-cingulate electrode that remained in NR1 throughout both seizures; (**ii**) an anterior cingulate electrode that remained NR2 in both; and (**iii**) a simultaneously recorded hippocampal electrode that transitioned from not-recruited to recruited during the seizure while the others remained unrecruited. Note the similarity of trajectories across seizures within electrode, despite dissimilar trajectories across electrodes within seizure.

Differences between groups were not explained by differing ictal durations (NR mean: 43 seconds, range: 18– 198; R mean: 37 seconds, range: 13–66), nor in the proportions of seizure types or mesial temporal versus lateral temporal onsets, although the proportion of extra-temporal onset zones was higher in NR (**Supplementary Table 2**; 48.5% (16/33) & 10.5% (2/19) in NR and R respectively, p = 0.014 chi-square). Differences were not due to patient-nor seizure-specific patterns: seizures from four patients (PT 4, 5, 8, 10) included simultaneous recordings across anatomical sites that were classified into different groups within the same seizure, and in three cases the same contacts were classified into different groups across seizures (PT 4, 11, 12; (**Fig. 4**; **Supplementary Table 2 & Supplementary Fig. 2**).

Hippocampal electrodes were evenly distributed between groups (NR = 14/33; R = 14/19), while the remainder of recordings were predominantly from cingulate (NR = 18/33; R = 3/19; **Supplementary Table 2**). For the 14 recruited hippocampal recordings, the SOZ was proximal: ipsilateral mesial temporal in 11 and ipsilateral neocortical temporal with later spread to hippocampus in 3. In contrast, for hippocampal recordings in NR, the SOZ was not ipsilateral mesial temporal in any, instead being ipsilateral, extratemporal neocortical for 8, contralateral mesial temporal for 2, and contralateral neocortical for 4 (**Supplementary Table 2**).

The NR group was further subdivided in the 3-cluster solution (**Fig. 5A**; “NR_1_” and “NR_2_”). Unlike the R/NR distinction, these sub-groups were not distinguishable by PLVHG (**Fig. 5Bv**; Bonferroni-Holm-corrected rank-sum p ∼1), nor line-length (a quantitative proxy for visual EEG assessment; p = 0.91). However, differences in measures reflecting population firing activity (HGA, FR, MU-E, and FWHM) distinguished the two (**Fig. 5;** p < 0.05 in each, Bonferroni-Holm-corrected rank-sum). In general, ictal firing patterns in NR_1_ more closely resembled recruited tissue than did those in NR_2_, though without the phase-locking to the dominant ictal rhythm indicative of local recruitment into seizure-driving tissue. Marked increases and decreases in firing rate during the seizure was observed in all groups. The heterogeneity in R differed from our prior neocortical studies, so we explored this further using single-unit analysis.

**Fig. 5.**
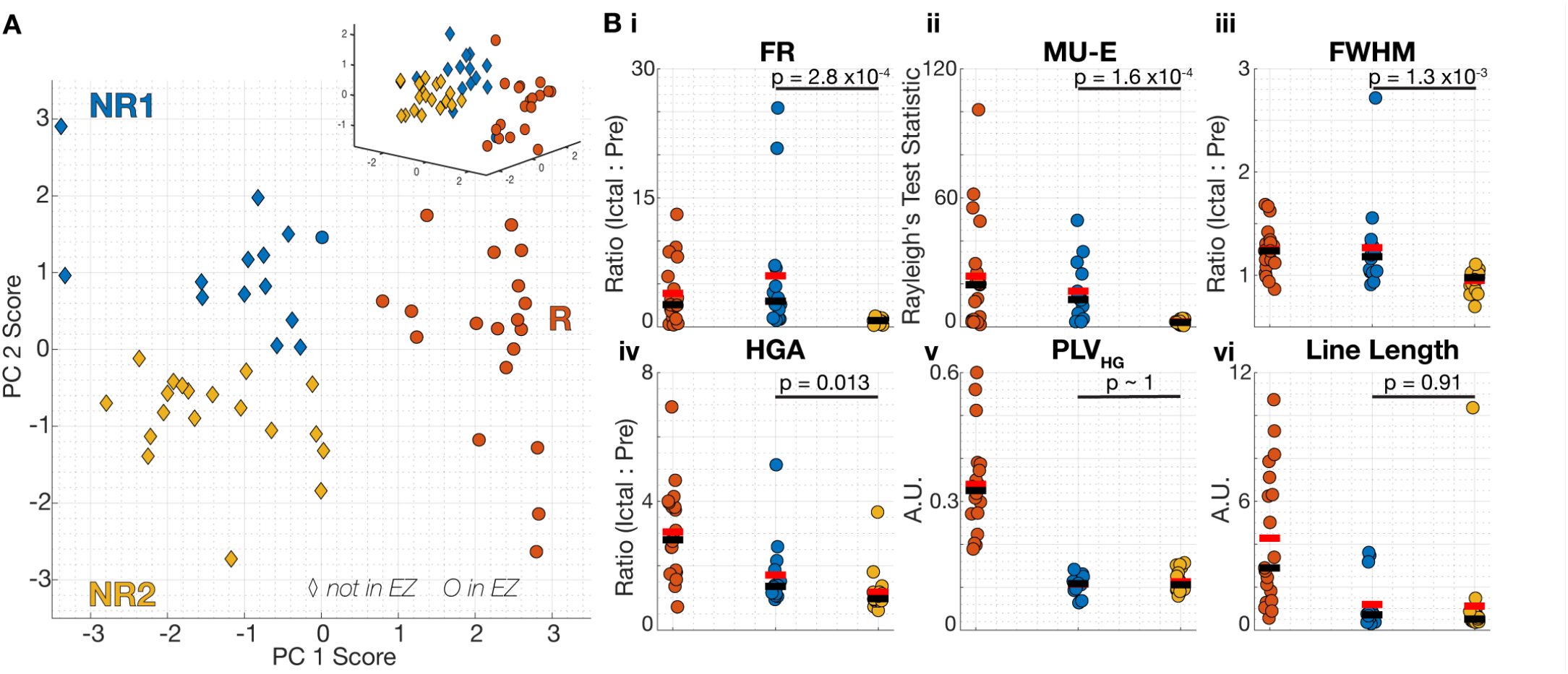
The 3-cluster solution reveals a subgroup of electrodes within the non-recruited group. **A**.Data plotted as in Fig. 3, but color coded to demonstrate the 3-cluster solution. In this solution the third cluster represents a subgroup of macro-micro pairs within the non-recruited group termed NR1 (blue) and NR2 (yellow). The recruited group (“R”) is the same in both the 2 and 3-cluster solution. **B**. Data plotted as per Fig. 3B but with subgroups NR1 & NR2 isolated. Only comparisons between the subgroups are made to compare ictal activity pattern. Compared to NR2, subgroup NR1 displayed significantly greater values in all metrics except for PLVHG and line-length. Subgroup average and median values are represented by the red and black bar, respectively.

### Single-unit analysis reveals cluster and cell-type differences in neuronal activity patterns

Individual single-units (*n* = 156 units; 34 seizures; 19 patients) were probabilistically subclassified as excitatory and inhibitory cells, based on waveshape (fast-spiking; FS interneurons) or subsequently via autocorrelation (RS interneurons; see Methods)^57^. Seventeen and 21 units (10.90% and 13.46%) were classified as FS and RS interneurons respectively at a confidence threshold of 50%; this classification was stable: restricting to a 95% confidence threshold reduced these numbers minimally (15 and 18 in total; 9.62% and 11.54%; **Supplementary Fig. 3**). A cutoff of 50% likelihood was therefore used for all categorical analyses by cell-type.

In contrast to the results for multi-unit firing, which includes all identifiable action potentials, ictal single-unit firing rates tended to decrease. Firing rate reductions were seen in 69.2% of single units, with 24.4% demonstrating decreases greater than 3 SD from pre-ictal firing rates (**Fig. 6A**). Firing rate increases exceeding pre-ictal baseline by 3 SD were seen in a minority (5.1%) of single units. No difference was found between recruited and non-recruited groups overall (n = 107 & 49 units respectively; p = 0.59, unpaired t-test) or by cell-type (p = 0.13 & 0.56; excitatory and inhibitory cells respectively; unpaired t-test). Overall, there was a trend towards larger decreases in firing rates of inhibitory compared to excitatory neurons during the seizure (**Fig. 6B**; p = 0.065; unpaired t-test). When separated by groups, however, the excitatory population was less impacted than inhibitory cells in the non-recruited group (Poisson-derived z-score mean ± SD: -0.79 ± 2.09 vs. −2.28 ± 3.77 respectively; p = 0.012, unpaired t-test). Meanwhile decreases were equivalent across cell-types in the recruited group (−1.36 ± 5.76 vs. −1.46 ± 2.51, inhibitory and excitatory cells respectively; p = 0.93, unpaired t-test). These changes were similar across contacts in SOZ versus those in regions of spread (−1.10 ± 2.83 vs. −1.38 ± 5.32 respectively; p = 0.220, rank-sum), however regions of spread showed higher variance (p = 0.020, Ansari-Bradley test). Within the non-recruited subgroups, interneuronal firing rate remained reduced in NR1 (p = 0.0058, unpaired t-test), while in NR2 both excitatory and inhibitory populations’ firing remained minimally altered from preictal rates (p = 0.68, unpaired t-test; **Supplementary Fig. 4**).

**Fig. 6.**
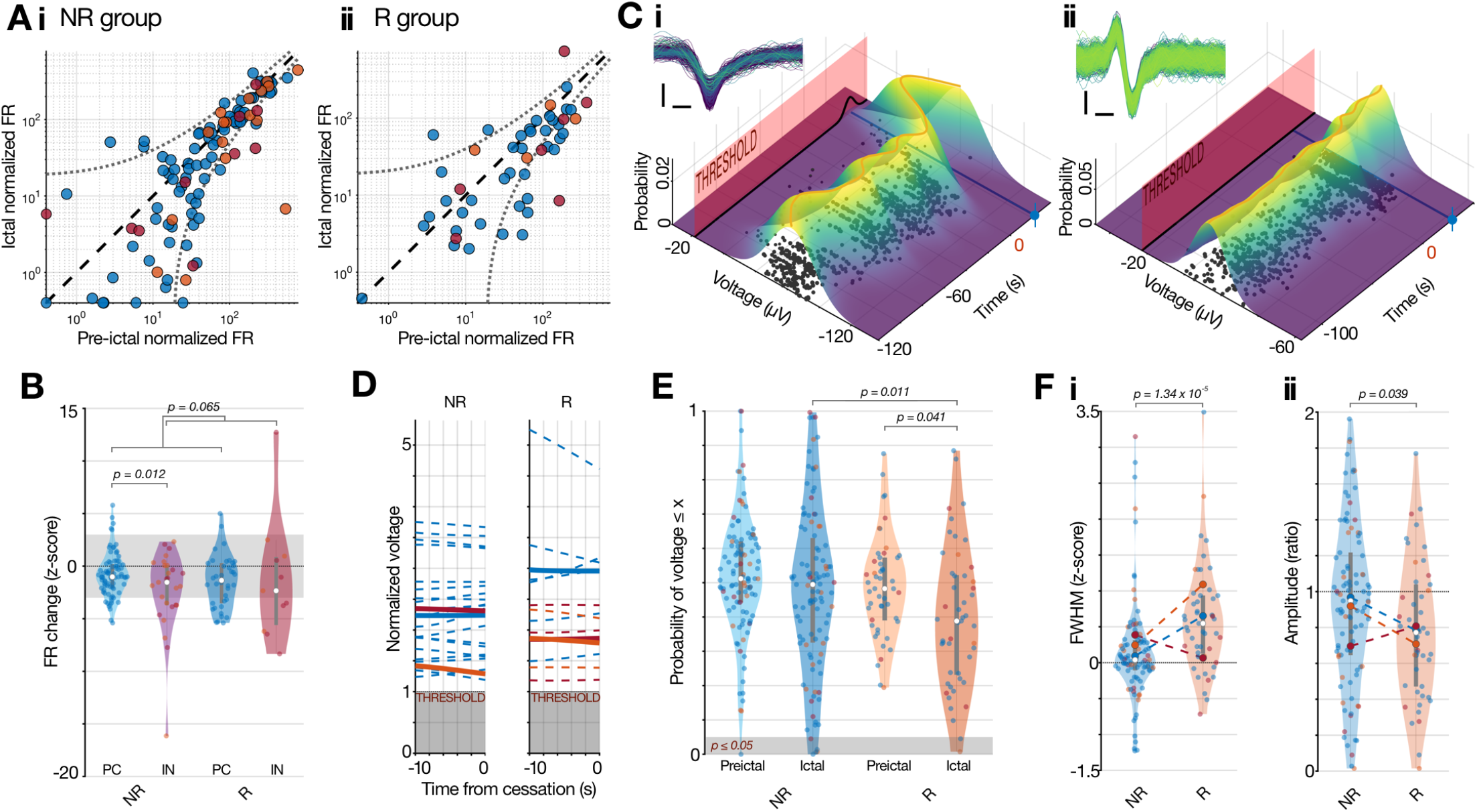
Single-unit ictal firing pattern analyses by putative cell-type. **A**.Pre-ictal vs. ictal normalized firing rates for putative pyramidal cells (blue), and two classes of interneurons (fast-spiking [red] and regular spiking [orange]—the same color-coding applies for other panels) in the non-recruited (**i**) and recruited (**ii**) groups. Dashed line represents equal firing rates across epochs, dotted lines show ± 3 S.D. firing rate changes as estimated by a Poisson distribution for that firing rate^65^. Dots on axes lines represent zero firing in that epoch. **B**. Violin plots of the ictal firing rates as calculated in A, z-scored by the Poisson-estimated S.D., grouped by excitatory and inhibitory putative cell-types. Each dot is a single-unit during one seizure (in NR: 80 pyramidal cells; 27 interneurons. In R: 38 pyramidal cells; 11 interneurons). Shaded region denotes ± 3 S.D. change from pre-ictal rates. **C**. Calculation of single-unit amplitude trajectories through time, showing an example of cessation of firing (blue line and marker) with increased chance of sub-threshold firing (**i**) and a cessation with stable action potentials (**ii**). To assess the trajectory of action potential amplitudes, a smoothing spline (λ = 0.001; orange line) was fitted to the voltage at detection (i.e. the spike trough value) for each spike through time for each unit, with the spikes weighted by their probability of being a true match to that neuron. The instantaneous shifted probabilistic distribution through time is shown in the heatmap. Threshold for spike detection with instantaneous probability of spikes from that neuron being subthreshold are shown as the red barrier and black line respectively. Black dots represent spike time and voltage at trough, and are scaled by confidence that spike came from its assigned neuron, with all spike waveforms shown inset. Scale bars: 0.2 ms, 50 μV (i) & 20 μV (ii); color scale showing time of spike. **D**. Trajectories from fitted spline in 10 s prior to cessation, normalized as multiples of threshold for detection for the given channel, by group and putative cell-type. **E**. Probability of the mean of the fitted spline (orange in C) in the pre-ictal and ictal epochs from the CDF of the pre-ictal distribution of voltages for each single-unit, by group and putative cell-type. Note that 0.5 represents the mean value, such that each value is the probability of getting that voltage or smaller from the original single-unit’s distribution. **F**. Waveform metrics for each single-unit, showing a population (**i**) increase in spike full-width at half maximum (FWHM) and (**ii**) decrease in amplitude in R, normalized to preictal values for each unit. Note the inverse relationship in the FS interneuron population across groups (red).

We next explored whether a subset of neurons ceasing firing entirely might explain the lack of overall increased firing rates associated with ictal invasion. Despite strict criteria for cessation (see Methods; **Fig. 6C**) and the permissive template-matching, 34 units (21.8%) ceased firing entirely prior to pre-termination. The proportion of inhibitory neurons that became silent was higher in R than NR (5/11, 45.5% vs. 2/27, 7.41% respectively; p = 0.0061, chi-square test). Excitatory cells also demonstrated cessation of firing, though similarly across groups (26.3% vs. 15.8% in NR and R respectively; p = 0.21, chi-square test). The majority of these excitatory cessations were in NR2 rather than NR_1_ (35.3% vs. 8.9%; p = 9.13 × 10^−4^, chi-square test; **Supplementary Fig. 4**).

To assess whether apparent firing cessations could be an artifact of spike detection methods, we tracked spike amplitude trajectories for each unit and determined the theoretical proportion of spikes that would be expected to be subthreshold for detection at any moment (**Fig. 6C**; see Methods). Trajectories prior to cessation were variable (**Fig. 6D**; 60.9% and 36.4% decreasing in amplitude in NR and R respectively; p = 0.18, chi-square test), although none had its mean amplitude reach threshold. The proportion of spikes from each unit that was likely subthreshold at the moment of cessation was similar across both groups, and not sufficient to explain the detected cessations as artifactual (NR: 5.03% ± 9.97% [median: 0.03%] vs. R: 3.71% ± 7.98% [median: 0.02%]; p = 0.94, rank-sum). To determine the impact of spike detection thresholds on firing rate calculations during the seizure, the same probabilistic method for the subthreshold proportion of spikes was calculated continuously, regardless of cessations (**Fig. 6E)**. The average proportion likely to be subthreshold was similarly low in both groups (NR: 3.01% ± 10.62% [median: 0.06%] vs. R: 2.67% ± 5.87% [median: 0.08%]; p = 0.85, rank-sum). Thus, neurons did not appear to stop firing due to depolarization block or to have undergone enough action potential waveform alterations to cause spike shapes to become undetectable.

### Ictal recruitment by cell-type and territory

To quantify seizure recruitment of individual neurons as demonstrated in previous neocortical studies^3^, we assessed waveform stability by cell-type in each group. Excitatory cells in the recruited group showed larger increases in spike duration during the seizure compared to the non-recruited group, reflecting the equivalent multi-unit increases (**Fig. 6Fi**; p = 9.0 × 10^−6^, rank-sum), but no difference was found in inhibitory neurons across groups (p = 0.36, rank-sum). On inspection, this apparent stability was comprised of dichotomous responses of the two putative types of inhibitory cell across the groups: RS interneurons trended in the same direction as the excitatory population (p = 0.047, unpaired t-test), while FS interneurons trended in the opposite direction, in keeping with intact feedforward inhibition mediated by PV^+^ interneurons. Similar trends existed for amplitudes (**Fig. 6Fii**). The larger changes in FS interneurons in the non-recruited group were dominated by subgroup NR1 (**Supplementary Fig. 4**), in keeping with intact feedforward inhibition mechanisms, however these were at chance levels with low numbers of observations (0.67 ± 1.41 vs. 0.04 ± 0.39; n = 6 & 4; p = 0.56).

Since a marked increase in firing followed by an abrupt decrease in FS interneurons, suggestive of depolarization block, has previously been associated with neocortical ictal invasion^4,30^, we sought to characterize the timing of FS interneuron firing rate alterations during the seizure. The aforementioned pattern was detected in only one of the seven FS interneurons in the recruited group (**Fig. 7A**; a transient increase >20 SD from pre-ictal levels based on a 5-second SD Gaussian convolution) while the remainder decreased firing without prior increases (**Fig. 7B**). In the non-recruited group, two of 10 FS interneurons transiently increased firing above 3 SD (both from the 6 FS interneurons in NR_1_), while the remainder of the 17 total FS cells in the study decreased firing rate without any prior increases (**Fig. 7C**). Assessing these transient increases over a range of detection parameters found FS interneuron firing rate increases lasted longest in NR1, followed by R, in contrast to other cell-types where increases were dominant in recruited tissue (**Supplementary Fig. 5**).

**Fig. 7.**
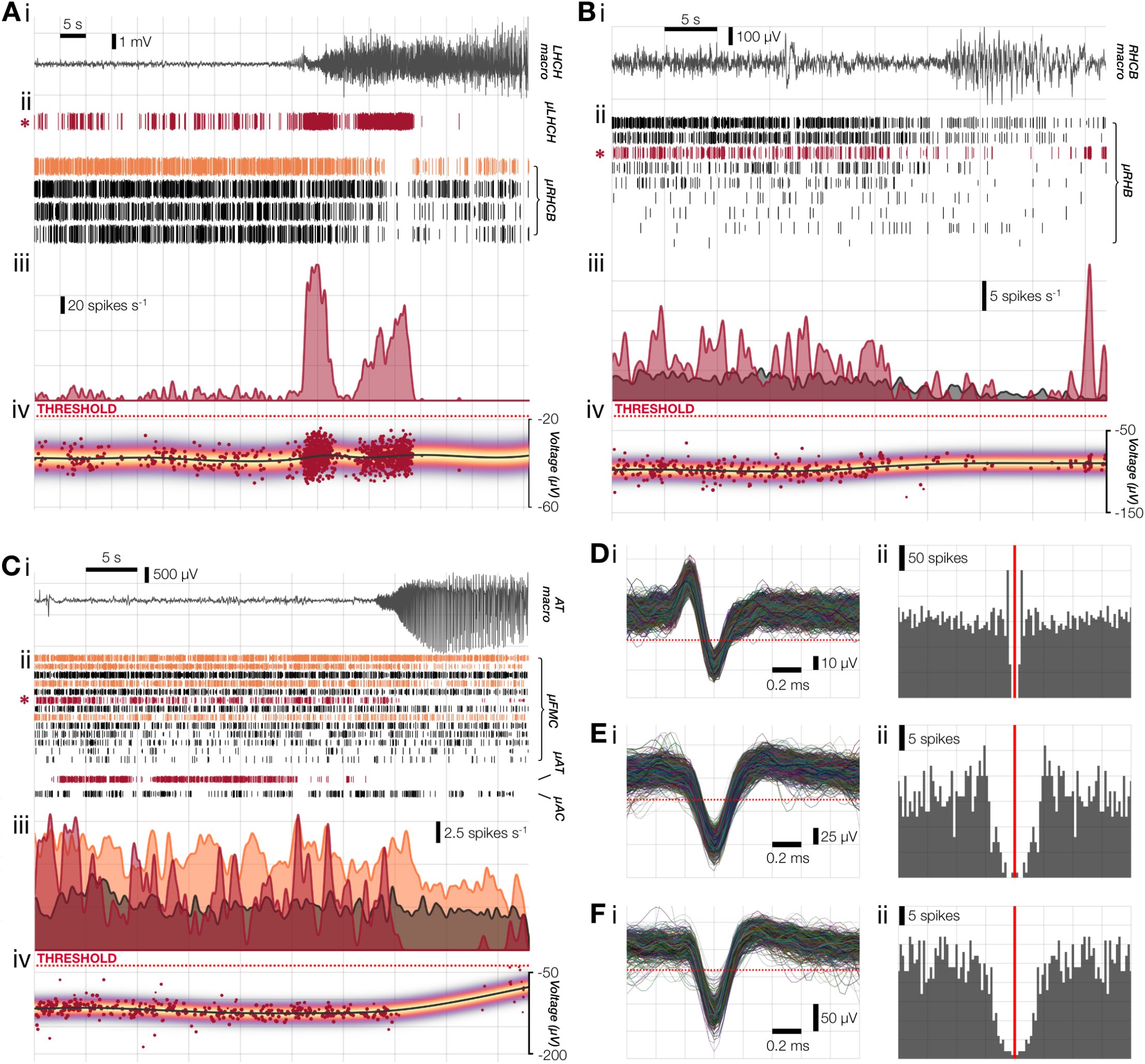
Example fast-spiking interneuron ictal firing patterns. **A**.A transient increase followed by cessation in the recruited group; **B**. A reduction of firing with no prior increase also in the recruited group; and **C**. A cessation with no prior increase in group NR1. In each: (**i**) LFP from the labeled macroelectrode; (**ii**) paired raster plot of all units from each microwire bundle, showing putative pyramidal cells (black), RS interneurons (orange) and FS interneurons (red). FS interneuron of interest is marked with an asterisk. Line height for each spike in the raster shows confidence each action potential arose from its assigned unit^3^; (**iii**) probabilistic firing rates for the marked FS interneuron along with the mean rates from other cells on the same microwire bundle (cell-type colors maintained from [ii]); (**iv**) voltage at detection for each spike from the marked FS interneuron (red dots, size shows confidence of match to that neuron), with fitted spline (black line), instantaneous probabilistic distribution through time (heatmap; see Fig. 6C), and threshold for detection (red dotted line). Note the stability and distance from threshold prior to each reduction or cessation of firing. **D-F**. For the example FS interneurons in A-C respectively: (**i**) Waveforms showing stability of waveshape and clearance from detection threshold (red dotted line); and (**ii**) autocorrelations over ± 100 ms.

Lastly, we sought to characterize changes in intrinsic firing patterns during seizures. The dissimilarity in the autocorrelations for each unit (**Fig. 8A&B**; “AC_dist_”; see Methods) showed a larger change in R than NR (0.22 ± 0.17 vs. 0.13 ± 0.09; p = 0.01, unpaired t-test; **Fig. 8C**). While AC_dist_ captures shifts in the temporal pattern of firing, it is ambiguous as to the specific direction of the alteration, so we examined both the average autocorrelations by cell-type (**Fig. 8D**) and the magnitude of *increases* in AC_dist_ (“AC_dist_+”; rather than absolute deviation) in order to characterize transitions towards burstiness. The recruited group showed more burstiness than the non-recruited group in both excitatory (AC_dist_+: 0.19 ± 0.20 vs. 0.09 ± 0.08; p = 0.015, unpaired t-test) and inhibitory cells (AC_dist_+: 0.25 ± 0.13 vs. 0.08 ± 0.10; p = 0.014, unpaired t-test), with inhibitory cells in the recruited group displaying a marked transition from cell-intrinsic steady-firing to burst-firing during the seizure (**Fig. 8D**).

**Fig. 8.**
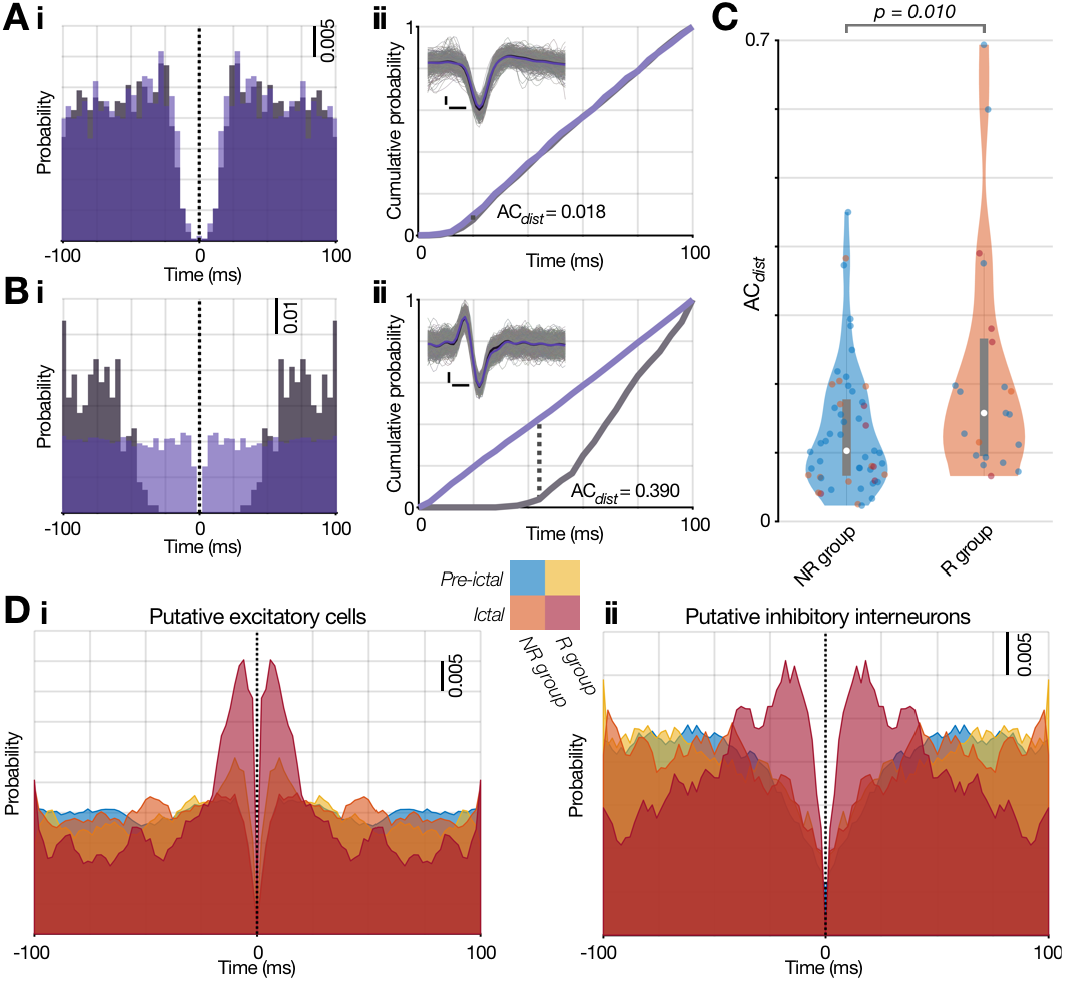
Alterations to cell-intrinsic firing properties in recruited tissue. **A & B**. (**i**) Example autocorrelograms from the pre-ictal (gray) and ictal (purple) epochs for two putative FS interneurons, with one showing stability in firing pattern (A) and the other transitioning to tonic firing during the seizure (B). (**ii**) Cumulative probability plots of the autocorrelograms in (i), showing ACdist calculation method, being the maximal difference between the two epochs’ normalized cumulative autocorrelations akin to K-S test. All waveforms are inset (gray) and mean from pre-ictal/ictal overlaid in black/purple, showing stability of spike shape. Scale bars: 0.2 ms, 20 μV (A) & 10 μV (B). **C**. Population ACdist values by group and cell-type. In order to derive meaningful autocorrelation patterns over the 100 ms window, this was limited to units with firing rates ≥ 0.2 spikes s^−1^ (i.e. at least 20 spikes contributing to the autocorrelation) **D**. Average autocorrelograms for each epoch and group, for putative (**i**) excitatory and (**ii**) inhibitory cells, showing the distinctive increase in burst firing (within 10 ms) for excitatory cells and shift from quintessential inhibitory interneuron firing pattern to a bursting pattern specific to the recruited group. Translucent colors as per key: blue & yellow for the pre-ictal epoch in groups R and NR respectively; orange & red for NR & R in the ictal epoch.

## Discussion

Seizure localization remains a significant challenge, limiting the efficacy of surgical management of refractory focal epilepsy. We have previously identified a critical division between neocortical regions that appear to be involved in seizures on EEG recordings: core territories that are “recruited” into a seizure, driving its propagation, versus penumbral tissue that is secondarily impacted by the ictal discharges but is not actively driving the seizure^1,6^. However, it is not clear how this dual-territory framework might apply to mesial temporal structures given the unique role of this region in temporal lobe epilepsy. Moreover, relating these territories to clinically accessible metrics is complex, as macro-contact signals are dominated by synaptic input to the region, rather than the local neuronal firing (the output)^66^. To address these questions, we utilized human multiscale recordings from limbic structures to identify modes of neural activity during seizures, linked to electroclinical metrics of site-specific seizure involvement.

### Three distinguishable seizure territories

The combined dataset of macro- and micro-electrode features demonstrated three distinct ictal activity patterns in limbic structures, with one group (R) displaying an increase in synchronous multi-unit firing entrained to the ictal rhythm and extracellular action potential waveform changes consistent with paroxysmal depolarization **(Fig. 3**). Both features have been observed during ictal recruitment in human and animal neocortical recordings^1,3,5^. The strong alignment of group R with the clinical assessment of the EZ and greater interictal HFO rate, along with predictions from previous studies of neocortical focal epilepsy in animal models and humans, supports our conclusion that the observed neuronal activity features specific to group R are representative of local ictal recruitment. Further, several instances of simultaneous recording of neural patterns indicative of recruitment and non-recruitment (group NR) provides new support for our previously described hypothesis of dual, co-existing but physically separate seizure territories: a relatively small ictal core is surrounded by penumbra that may show the EEG signature of a seizure but in which neural firing is restrained by intact inhibition^1,6,52,67^.

While all analyses were performed blind to the clinical assessment of seizure onset, of 20 total macro-micro pairs interpreted to be located in EZ, 19 were in group R. This accuracy is rather higher than is seen in neocortex, where the clinically determined EZ overestimates the spatial extent of tissue recruitment^1,43,52^. Given the prevalence of hippocampal recordings in this group (74%), one possible explanation is the ease of detecting seizure activity with sEEG in hippocampal structures given its unique cytoarchitecture and the relatively dense hippocampal electrode coverage. An alternative explanation might be that hippocampus is highly susceptible to ictal recruitment compared to neocortex, withstanding the synaptic barrages for less time, thereby resulting in minimal time between the first EEG signatures of seizure activity and subsequent recruitment of local neurons. Such region-specific differences of ictal dynamics would be in keeping with *in vitro* observations of differing neocortical and hippocampal ictal patterns in animals^68^ and in varying seizure susceptibility and ictal patterns in resected human tissue^69^. We also detected a subcluster within the non-recruited group (NR1), in which EEG-based metrics did not reveal ictal recruitment, but single and multi-unit metrics more closely resembled the pattern seen in the recruited group, suggesting feedforward effects from the seizure at these sites. This is consistent both with the neuronal activity expected in penumbral territories receiving synaptic barrages from the seizure core^1,5,70^ and with our recently published observations of neocortical ictal recruitment operating at the level of single neurons^3^. Such sites may represent a strongly connected node involved in the propagation of the seizure itself^71,72^ by amplifying the excitatory barrages experienced at the distant site and increasing the possibility of secondary recruitment at that site. Another possibility is that incipient recruitment could result from locally impaired inhibition. A rodent model of acute neocortical seizures has demonstrated that focal seizures could generate distant secondary foci in the absence of pathologically reinforced structural alterations as a result of local inhibitory breakdown, which can appear as coordinated network activity^8^. In either scenario, subgroup NR_1_ may be framed as representing secondary regions that have the potential to transition into an ictal state, while NR_2_ is dominated by territories less-impacted by the ongoing seizure.

An intriguing question is whether NR1 sites have increased potential to become dominant seizure foci following resection/disconnection of the primary focus. Consider the post-surgical outcomes of patients 1 and 13, who both had extratemporal SOZs with hippocampal recordings belonging to subgroup NR_1_: PT 13 had an Engel 3A outcome at 16.5 months following an insular resection, and PT 1 had a frontal lobe resection with an Engel 1A outcome at 19 months. However, PT1 went on to have a second surgical evaluation in light of non-specific sensory auras that later arose, although with no evidence of hippocampal involvement (**Supplementary Table 1**). Future studies may elucidate if sites with NR1 activity patterns might predict post-surgical failure.

Of the examined neuronal activity patterns, high-gamma entrainment to the ictal rhythm, as indexed by PLV_HG_ obtained from the macroelectrode, was the only metric capable of distinguishing the two ictal territories at the level of an individual seizure and recording site. This finding is consistent with previous studies validating EEG-derived metrics that simultaneously assess both high- and low-frequency information, including the related phase-locked high-gamma metric^43,52^ and the epileptogenicity index, which incorporates line-length but does not consider phase-locking^73^. Even with implanted depth electrodes, the ictal wavefront (a band of tonic firing at the leading edge of an advancing seizure), is unfortunately largely invisible to clinical EEG recordings likely due to abolishing the periodic firing that is a basic requirement for generating oscillatory EEG features^1,6,35,40,43^. High-gamma activity phase-locked to the underlying ictal rhythm, however, has been shown to increase sharply following passage of the ictal wavefront but not at sites that did not demonstrate an ictal wavefront^1,43^. PLV_HG_, therefore, is a practical method of identifying ictal recruitment on a per seizure basis. Conversely, measures such as multi-unit firing rate or high-gamma activity, while demonstrating measurable differences between groups, may not reliably categorize the recruitment status of an individual channel’s recording.

### Heterogenous single-unit changes during seizures

While the anticipated difference in multi-unit firing between recruited and non-recruited groups was found, the high degree of heterogeneity of firing within both groups differed sharply from our prior neocortical findings^1,3,5,35,43^. Although heterogeneity in ictal firing has previously been reported in human recordings^17,18,20^, we had hypothesized that heterogeneous firing would be much less prominent in recruited brain areas, on the basis of our prior observations in neocortical microelectrode human recordings^1,3^. We explored single-unit firing in detail using our recently developed template-matching method, which was designed to avoid confounding firing rate assessment due to changes in noise level and action potential waveform shape^3,4^.

Although a natural decrease in firing rate due to the well-known hyperpolarization that follows epileptiform discharges may occur, average firing rates remained high in our prior analyses of neocortical recordings^1,3,4,35^. In contrast, across all 156 single-units in this study, we found that a high proportion (69%) of all cell-types demonstrated decreases in ictal firing rates (**Fig. 6A-B**), with some ceasing firing altogether. This prominent decrease appears to contradict the simultaneous findings of increased high-gamma amplitude and PLV_HG_, recorded from the nearby macroelectrode, due to the association of ECoG-recorded high-gamma activity with intense multi-unit firing^35,36^. One possible explanation is that single-unit detection is inherently biased towards high firing rate neurons, and these may be more likely to decrease their firing rate than neurons that fire less frequently.

### Firing patterns of fast-spiking interneurons

Recent work has uncovered evidence of marked FS interneuron firing rate increases at seizure onset, with a proposed “excitatory rebound” mechanism contributing to ictogenesis^28,74^. A subsequent study of human microelectrode data reported finding evidence of increased interneuron firing early in the seizure in a group of 9 patients with low-voltage fast onset patterns^30^. In our dataset, one out of 17 FS interneurons, an interneuron from the recruited group, demonstrated a striking activity pattern of transient increases in firing followed by cessation of firing (**Fig. 7A**). This FS interneuron firing pattern has been ascribed to a rapidly increasing excitatory drive which may result in simultaneous weakening or even reversal of the GABA current in the postsynaptic cell due to intracellular chloride loading^4,75–79^. Under this scenario, a dramatic increase in pyramidal cell firing is expected following the sharp increase in FS interneuron activity, as we have demonstrated in neocortical human recordings^4^. This increase was not observed in our data in any group (**Supplementary Fig. 5**), though it should be noted that in this particular example of interneuronal activity, there were no pyramidal cells captured at the same electrode. Nonetheless, the interneuronal activity seen here is consistent with observations in a pilocarpine-mouse model where interneuronal inactivation during the seizure was common, while pre-ictal activation or inactivation was heterogeneous and dependent on hippocampal subfield^27^.

FS interneuron depolarization block is another mechanism that has been proposed to underlie the interictal-ictal transition and subsequent recruitment of post-synaptic pyramidal cells^25,77,80,81^. Yet, we did not find definitive evidence of depolarization block in the sampled mesial structures to explain either the cessation or reduction in FS interneuron firing, despite the clear presence of a highly excitatory state: single-unit trajectories in these recordings were generally stable and clear of the detection threshold during the seizure (meaning that drop-out was not explained by small fluctuations in the shape of the unit; **Fig. 6E**), and the majority of FS interneuron firing rate reductions occurred without a prior increase (**Fig. 7B-C**).

For the limbic seizures in this study, these findings suggest that the posited weakening of local inhibition, while occurring sporadically, is unlikely to be a major driver of hippocampal seizures. Nevertheless, there is some evidence for feedforward inhibition and its subsequent ictal impairment. The largest transient increases in FS interneuron firing, as well as maximal waveshape changes of the FS population prior to reduction in firing, occurred in the group that remained unrecruited, though had multi-unit evidence of oncoming ictal activity (NR_1_; **Supplementary Figs. 4&5**), and once recruited, interneurons showed perturbed, bursting firing patterns (**Fig. 8**). As such, inhibition is likely compromised to a degree, but may not be the complete inhibitory failure that has been documented in slice studies or neocortical recordings.

We note that, as is common in the patient population undergoing invasive monitoring of mesial temporal structures^82^, none of the patients in this study had mesial temporal sclerosis on preoperative imaging or tissue pathology (**Supplementary Table 1**). It may be that the non-sclerotic hippocampus, while readily becoming excitable, is less susceptible to complete inhibitory collapse compared to neocortex as a result of its unique cytoarchitectural structure. We propose that the predominance of single-unit firing rate reductions provides evidence for at least moderately maintained inhibition in the hippocampus for the majority of recordings in the recruited group, i.e. that if inhibitory failure does occur in the hippocampus during seizures, it is not a common phenomenon in this study population. This suggests a possible alternative scenario in which the hippocampus, while clearly in a high excitability, rhythmic activity state, may not have been completely recruited, instead amplifying seizure activity driven from a nearby source but not itself primarily driving seizure activity. Similar decreases in hippocampal interneuron firing were reported in 7 patients with known neocortical onsets and suspected mesial temporal spread^33^. This raises the interesting question of whether complete vs. partial hippocampal recruitment may have bearing on whether hippocampal removal can successfully prevent seizures, and whether clinically accessible information can be used to differentiate these two activity states.

## Funding

The authors acknowledge funding from NIH R01 NS084142 (CAS), NIH R01 NS110669 (CAS), and CRCNS R01 NS095368 (CAS).

## Competing interests

GMM is an investigator and on the publication committee for the “Stereotactic Laser Ablation in Temporal Epilepsy” (SLATE) trial funded by Medtronic, plc. None of the patients in this study were enrolled in the SLATE trial. The other authors report no competing interests.

## Supplementary material

Supplementary material is available online.

## Notes

### Author Declarations

The IRB of Columbia University gave ethical approval for this work

