## Supplementary material for "Cell-type specific and multiscale dynamics of human focal seizures in limbic structures"

**Running title**: LIMBIC STRUCTURE FOCAL SEIZURE DYNAMICS

Contents:

- Supplementary Figures 1–5
- Supplementary Tables 1 & 2

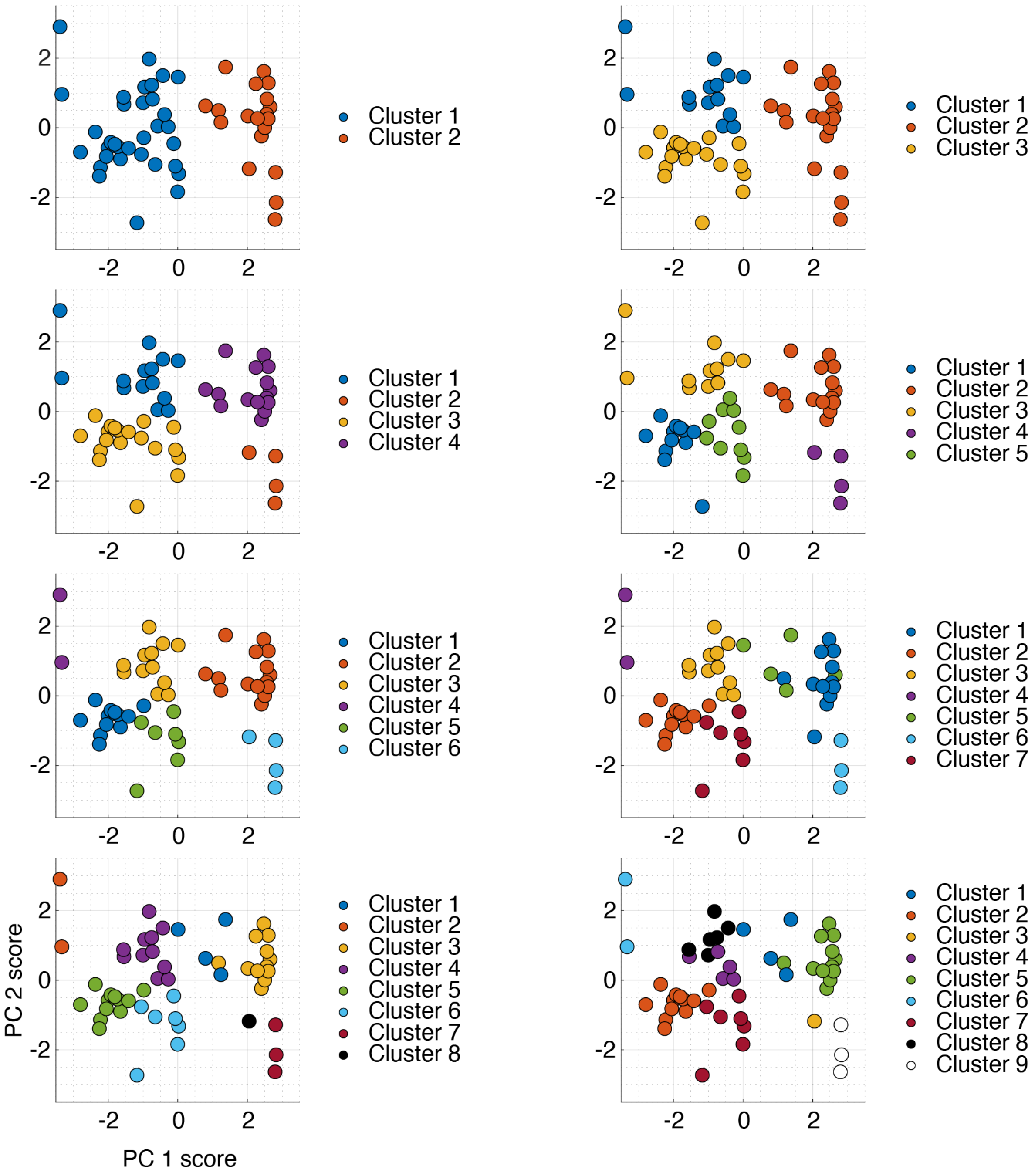


**Supplementary Figure 1. K-means cluster analysis of ictal neuronal activity metrics.** Projections onto the first two principal component scores for each $k$-means cluster solutions where $1 < k \leq 9$. Note that solutions with $k > 7$ produce one-member clusters and thus were not evaluated further.

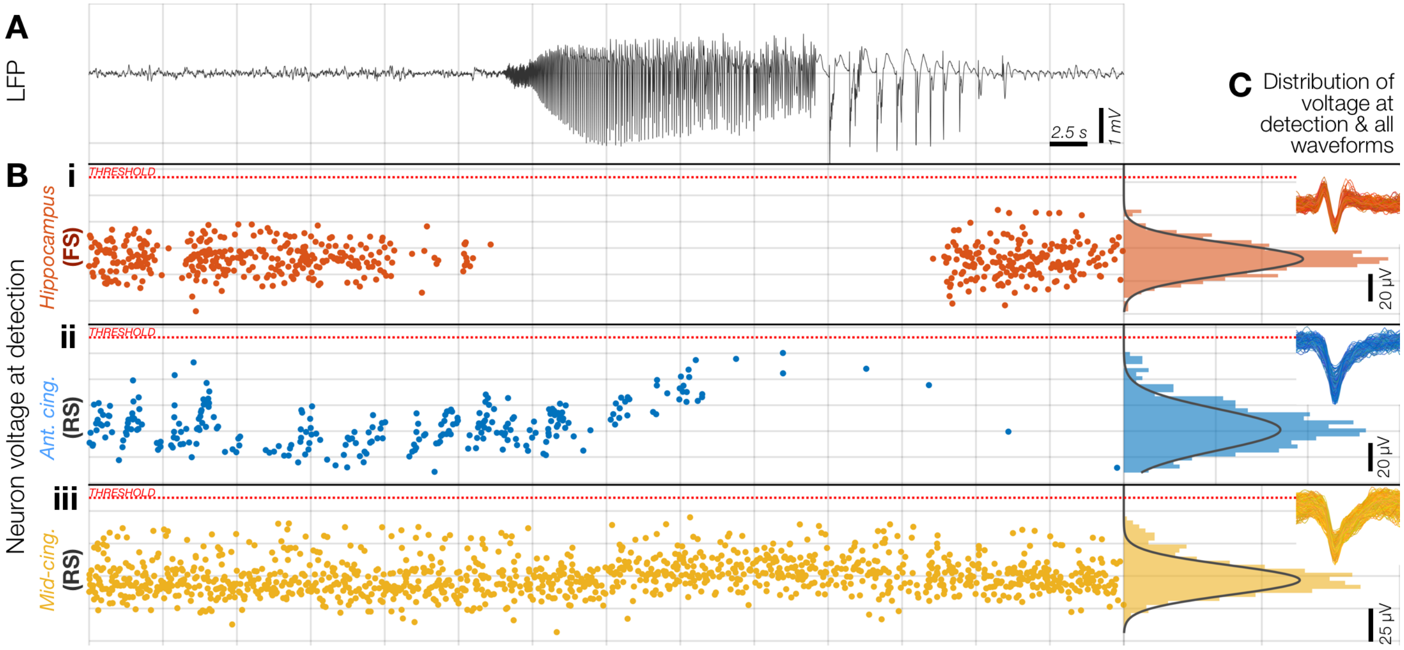


**Supplementary Figure 2. Examples of single-unit firing rate changes across the ictal transition observed in the three identified ictal patterns during a single seizure. A.** LFP of a seizure captured in the hippocampus. **B.** Voltages of detected action potential troughs plotted against time with respect to the LFP in A and the threshold for detection (red dotted line) for (**i**) a fast-spiking interneuron in the hippocampus, and (**ii**–**iii**) putative pyramidal cells in the anterior and mid-cingulate respectively. **C.** Histograms of all detected action potential voltages shown in B with Gaussian fits to estimate percentage of missed detections and all detected waveforms for each unit, inset. Note the stability of each single-unit's waveform and the low probability of spikes being below the detection threshold: the observed cessation or decrease in firing is unlikely to be a result of spike detection methods. Group color coding from the main text is maintained (orange = "R" group, blue = "$NR_1$" group, yellow = "$NR_2$" group), showing the sudden cessation of firing at seizure onset without any preceding firing rate changes in R (i), decreasing firing during the ictal transition, suggestive of intact feed-forward inhibition in $NR_1$ (ii), and a neuron that appears to be unaffected by the ongoing seizure activity in the hippocampus ($NR_2$; iii).

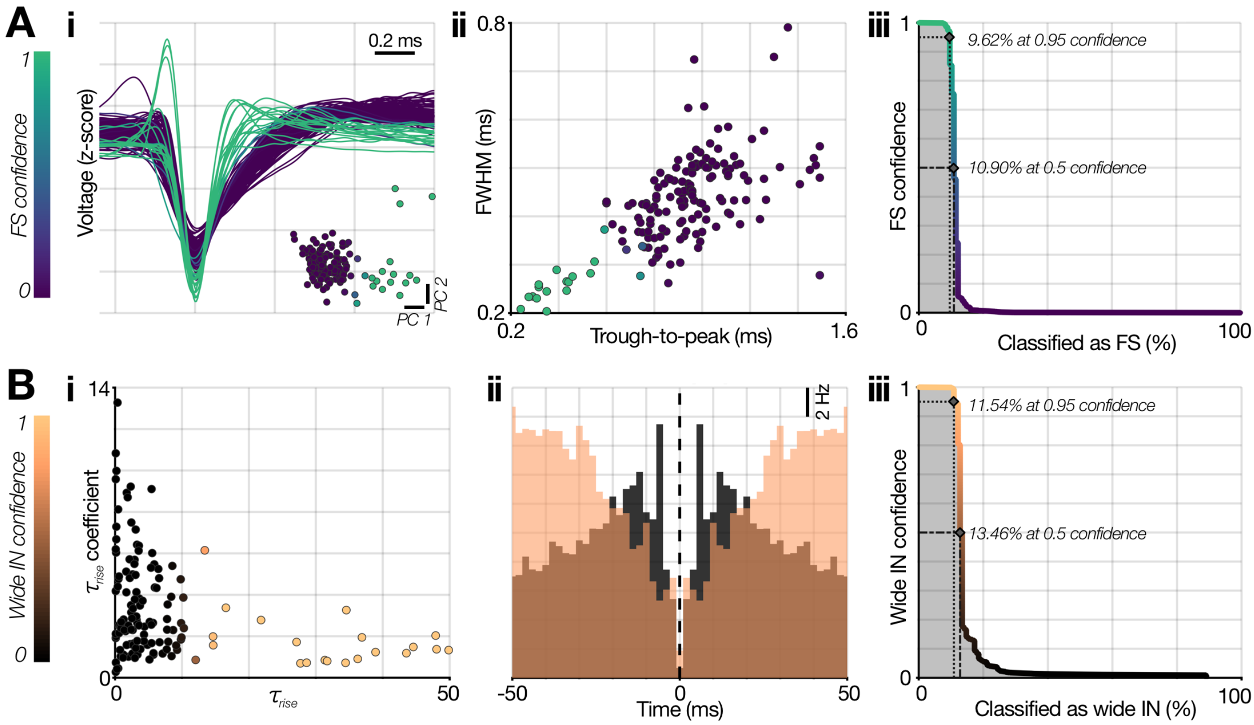


**Supplementary Figure 3. Cell-type subclassification of single-units. A.** Probabilistic subclassification of fast-spiking (FS) interneurons. (**i**) Mean wideband waveforms from each single-unit, color-coded by its confidence of being classified as a FS interneuron based on a 2-component Gaussian mixture model fitted to these waveforms' scores in PC space (inset; color-scale maintained throughout A). (**ii**) Confidences from this model showed the expected differences in spike duration in the putative FS interneuron population, and classification proved stable, with similar percentages of units being classified as FS interneurons across a wide range of confidence cutoffs (**iii**). **B.** Probabilistic subclassification of RS interneurons. (**i**) The autocorrelograms for each unit that was not classified as a putative FS cell were fitted with a set of exponentials to calculate $\tau_{rise}$ (Petersen et al., 2020; see Methods), which were then fitted with a 2-component Gaussian mixture model to derive confidences of each unit being a non-FS interneuron ("RS IN"; black to copper color scale; maintained throughout B). (**ii**) Mean autocorrelograms for wide single-units with > 50% and ≤ 50% confidence of being a RS IN (copper and black respectively). (**iii**) As for putative FS interneuron classification, the probabilistic classification of RS INs proved stable, with similar percentages of units being classified across a wide range of confidence cutoffs.

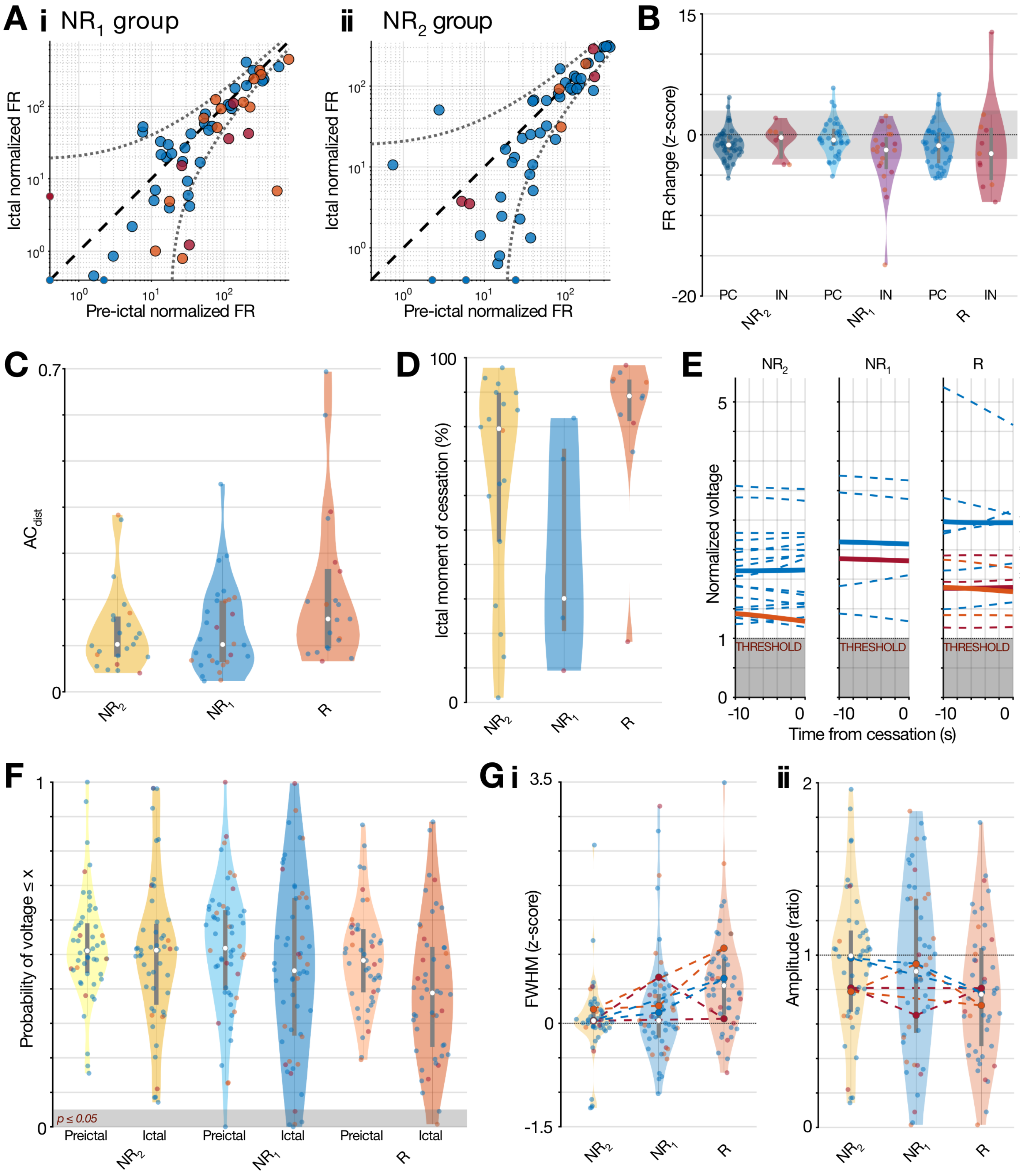


**Supplementary Figure 4. Single-unit metrics for the 3-cluster solution. A, B.** Normalized firing rates for groups $NR_1$ and $NR_2$, as per Fig. 5A, B. **C.** $AC_{dist}$ for all three groups as per Fig. 7C. **D.** Normalized ictal timing for each cessation across groups. **E.** Voltage trajectory for each cessation as per Fig. 5D. **F.** Probability of obtaining a voltage at least as low as observed from the original CDFs for each unit's Gaussian fit, with respect to group and epoch, as per Fig. 5E. **G.** Waveform metrics for each single-unit across all three groups showing (**i**) spike full-width at half maximum, and (**ii**) spike amplitude, as per Fig. 5F.

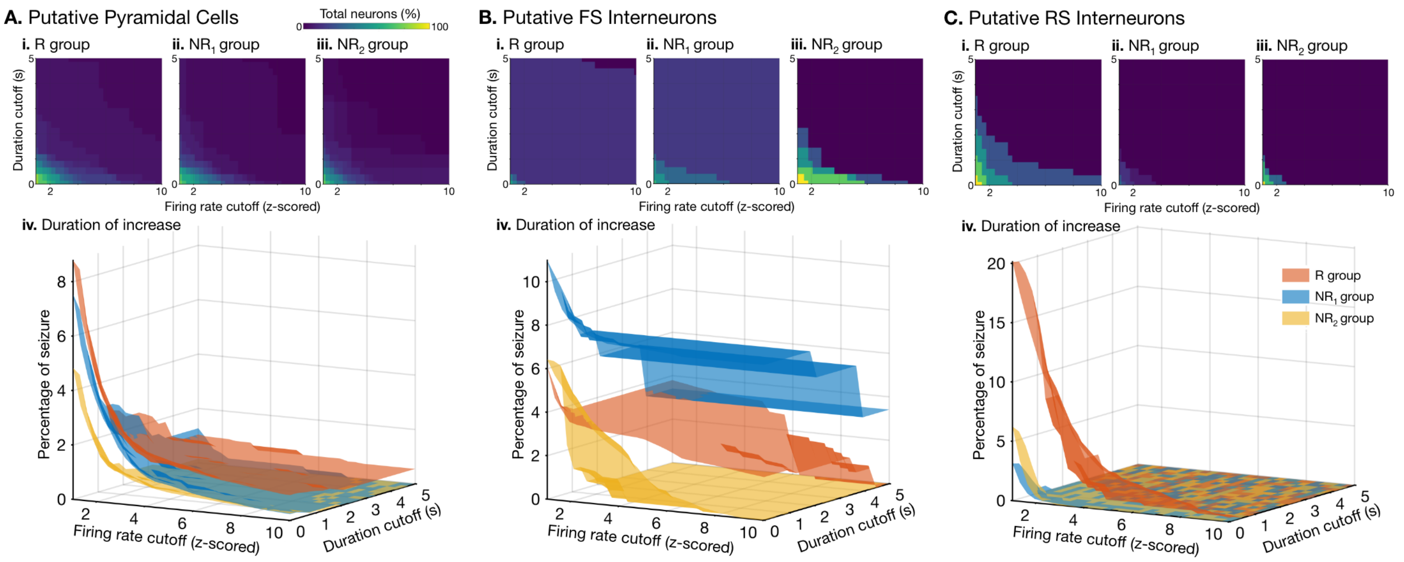


**Supplementary Figure 5. Transient firing rate increases by cell-type and group. A–C.** Firing rate increases for putative pyramidal cells, FS interneurons and RS interneurons respectively. In each, the percentage of units (color scale) that reached at least a given SD increase (z-score, x-axis) and duration (seconds, y-axis) for groups R, $NR_1$ and $NR_2$ in (**i–iii**) respectively. (**iv**) the duration of each increase at the given cutoffs (x- and y-axes) as a percentage of the seizure duration (z-axis) for each group (color), revealing the largest increases in the recruited group for putative excitatory cells (A) and RS interneurons (C), but in the incipient recruitment group (blue, $NR_1$) for the fast-spiking interneurons (B).

| PT | Age Range | Sex | # Seizures Analyzed | Lesional | MRI Findings (if applicable) | Surgical Pathology (if applicable) | Intervention | Engel Outcome | Follow Up Time (months) |
|---|---|---|---|---|---|---|---|---|---|
| 1 | 30–34 | F | 2 | Yes | Punctate foci of susceptibility in high medial left frontal and posterior right temporal regions. Could represent foci of calcification from neurocysticercosis | Fibrotic, calcified nodules | L. Frontal lobe resection | 1B | 18.1 |
| 2 | 25–29 | F | 1 | No | - | - | LITT: R. Amygdala-Hippocampus | 3A | 24.7 |
| 3 | 55–59 | F | 1 | No | - | - | R. Temporal lobectomy | 1A | 3.5 |
| 4 | 18–24 | M | 2 | No | - | - | R. Temporal lobe and insular resection | 1D | 29.1 |
| 5 | 18–24 | F | 3 | No | - | - | LITT: L. Amygdala-Hippocampus | 1D | 32.7 |
| 6 | 18–24 | M | 1 | No | - | - | R. Temporal lobectomy | 3A | 18.9 |
| 7 | 18–24 | F | 1 | No | - | - | LITT: R. Mesial temporal | 1A | 20.7 |
| 8 | 40–44 | M | 2 | No | - | - | L. Frontal lobe resection | 1A | 19.0 |
| 9 | 25–29 | F | 1 | Yes | Prior right lateral temporal lobe resection for choroid plexus carcinoma. No evidence of recurrence. Right anteromedial FLAIR hyperintensities likely postsurgical encephalomalacia and gliosis rather than mesial temporal sclerosis. | Moderate astrogliosis and microgliosis | R. Temporal lobectomy | 1A | 19.3 |
| 10 | 18–24 | F | 1 | No | - | - | RNS | 3A | 13 |
| 11 | 35–39 | M | 3 | No | - | - | None | N/A | N/A |
| 12 | 30–34 | M | 3 | No | - | - | RNS | 2B | 29.9 |
| 13 | 30–34 | M | 1 | No | - | - | R. Insular resection | 3A | 16.5 |
| 14 | 18–24 | F | 1 | No | - | - | R. Lateral temporal lobe resection | 1A | 27.8 |
| 15 | 35–39 | M | 3 | No | - | - | RNS | 4B | 12.3 |
| 16 | 45–49 | F | 3 | No | - | - | L. Medial frontal lobe resection | 1D | 15.2 |
| 17 | 50–54 | F | 1 | No | - | - | RNS | 4B | 13.6 |
| 18 | 20–24 | M | 2 | Yes | No evidence of residual/recurrent lesions around prior resection cavity for ganglioma. No other structural abnormalities identified | No evidence of abnormal tissue in hippocampus; low grade glioma along prior ganglioma resection cavity in inferior temporal gyrus | R. Anterior temporal lobectomy | 4A | 12.0 |
| 19 | 30–34 | M | 2 | No | - | - | No data | N/A | N/A |

**Supplementary Table 1. Patient demographics**

| PT | Seizure | Type | SOZ | # of Macro-Micro Pairs Included in Study | | | Total pairs per seizure |
|---|---|---|---|---|---|---|---|
| | | | | $NR_1$ group | $NR_2$ group | R group | |
| 1 | 1 | FTBTC | L. Lateral Frontal | Left Hippocampus | | | 1 |
| | 2 | FTBTC | L. Lateral Frontal | Left Hippocampus | | | 1 |
| 2 | 3 | Focal Unaware | L. Mesial Temporal | | | Left Hippocampus | 1 |
| 3 | 4 | Subclinical | R. Lateral Temporal | | | Right Hippocampus | 1 |
| 4 | 5 | Focal Unaware | R. Lateral Temporal | Right SMA | Right ant. Cingulate | | 2 |
| | 6 | FTBTC | R. Lateral Temporal | | | Right SMA<br>Right ant. Cingulate | 2 |
| 5 | 7 | Focal Unaware | L. Anteromedial Temporal | Left mid Cingulate | Left ant. Cingulate | Left Hippocampus 1 | 3 |
| | 8 | Focal Unaware | L. Anteromedial Temporal | Left mid Cingulate | Left ant. Cingulate | Left Hippocampus 1<br>Left Hippocampus 2 | 4 |
| | 9 | Focal Unaware | L. Anteromedial Temporal | Left mid Cingulate | Left ant. Cingulate | Left Hippocampus 1<br>Left Hippocampus 2 | 4 |
| 6 | 10 | FTBTC | R. Anterolateral Temporal | Right ant. Cingulate | | | 1 |
| 7 | 11 | Focal Unaware | R. Mesial Temporal | | | Right Amygdala | 1 |
| 8 | 12 | Focal Unaware | L. Lateral Frontal | Left mid Cingulate | Left Hippocampus | | 2 |
| | 13 | Focal Unaware | L. Lateral Frontal | Left mid Cingulate | Left Hippocampus | | 2 |
| 9 | 14 | Focal Unaware | R. Lateral Temporal | | | Right Hippocampus 1<br>Right Hippocampus 2 | 2 |
| 10 | 15 | Focal Aware | L. Mesial Temporal | Right Hippocampus | | Left Hippocampus | 2 |
| 11 | 16 | FTBTC | R. Mesial Temporal | | Right mid Cingulate | | 1 |
| | 17 | FTBTC | R. Mesial Temporal | | Right mid Cingulate | | 1 |
| | 18 | FTBTC | L. Orbitofrontal | Right mid Cingulate | | | 1 |
| 12 | 19 | Focal Aware | L. Mesial Temporal | Right mid Cingulate | | | 1 |
| | 20 | Focal Aware | L. Mesial Temporal | | Right mid Cingulate | | 1 |
| | 21 | Focal Aware | L. Mesial Temporal | | Right Hippocampus | | 1 |
| 13 | 22 | FTBTC | R. Frontal | Left Hippocampus | | | 1 |
| 14 | 23 | Focal Unaware | R. Lateral Temporal | | Right Hippocampus | | 1 |
| 15 | 24 | Subclinical | R. Insula + Somatosensory Cortex | | Left Hippocampus<br>Right Hippocampus<br>Right ant. Cingulate | | 3 |
| | 25 | Subclinical | R. Insula + Somatosensory Cortex | | Left Hippocampus<br>Right Hippocampus | | 2 |
| | 26 | Subclinical | R. Insula + Somatosensory Cortex | | Left Hippocampus<br>Right Hippocampus | | 2 |
| 16 | 27 | FTBTC | L. Cingulate | | Right ant. Cingulate | | 1 |
| | 28 | FTBTC | L. Cingulate | | | Left ant. Cingulate | 1 |
| | 29 | FTBTC | L. Cingulate | | | Left ant. Cingulate | 1 |
| 17 | 30 | Focal Unaware | L. Mesial Temporal | Right ant. Cingulate | | | 1 |
| 18 | 31 | Focal Unaware | R. Mesial Temporal | | | Right Hippocampus | 1 |
| | 32 | Focal Unaware | R. Mesial Temporal | | | Right Hippocampus | 1 |
| 19 | 33 | Focal Aware | Bilateral Mesial Temporal | | | Left Hippocampus | 1 |
| | 34 | Focal Aware | Bilateral Mesial Temporal | | | Left Hippocampus | 1 |

**Supplementary Table 2. Seizure and macro-micro pair information.** The number and type of seizures recorded per patient, including the clinically determined seizure onset zone (SOZ) for each seizure. Each recording macro-micro pair is listed by anatomical location and group identity from cluster analysis. FTBTC = focal to bilateral tonic-clonic. SOZ comparisons between the non-recruited (NR) group vs. recruited (R) group: mesial temporal 0.39 (13/33) vs. 0.63 (12/19), $p = 0.17$ chi-square; lateral temporal 0.12 (4/33) vs. 0.26 (5/19), $p = 0.35$ chi-square; extratemporal 0.48 (16/33) vs. 0.10 (2/19), $p = 0.014$ chi-square. Seizure type comparisons NR vs. R: FTBTC 0.24 (8/33) vs. 0.21 (4/19), $p = 0.93$ chi-square; focal unaware 0.42 (14/33) vs. 0.58 (11/19), $p = 0.43$ chi-square; focal aware 0.12 (4/33) vs. 0.16 (3/19), $p = 0.96$ chi-square; subclinical .21 (7/33) vs. 0.05 (1/19), $p = 0.25$ chi-square. SOZ comparisons Subgroup $NR_1$ vs. Subgroup $NR_2$: mesial temporal 0.43 (6/14) vs. 0.37 (7/19), $p = 0.99$ chi-square; lateral temporal 0.14 (2/14) vs. 0.11 (2/19), $p = 0.83$ chi-square; extratemporal 0.43 (6/14) vs. 0.53 (10/19), $p = 0.84$ chi-square. Seizure type comparisons Subgroup $NR_1$ vs. Subgroup $NR_2$: FTBTC recordings 0.36 (5/14) vs. 0.16 (3/19), $p = 0.36$ chi-square; focal unaware 0.5 (7/14) vs. 0.36 (7/19), $p = 0.69$ chi-square; focal aware 0.14 (2/14) vs. 0.11 (2/19), $p = 0.83$ chi-square; subclinical 0 vs. 0.37 (7/19).
